# Clonal Haematopoiesis Tracks Alzheimer’s-like Cortical Atrophy in Community-Dwelling Older Adults

**DOI:** 10.64898/2026.09.10.26362681

**Authors:** Y. Suwanlikit, S.J.C. Crofts, J.E. Moodie, R.E. Marioni, N.A. Robertson, S.H. Tan, P. Vivithanaporn, M.E. Bastin, J.M. Wardlaw, K. Kirschner, S.R. Cox, T. Chandra

## Abstract

**Background:** Clonal haematopoiesis (CH), the age-related expansion of mutated blood stem cells, is detectable in around 10% of people in their seventies, rises steeply with age, and is associated with several diseases. Its relationship with neurodegeneration remains contentious, with conflicting evidence for both protective and detrimental effects on Alzheimer’s disease (AD).

**Methods:** Here, we applied COMET, a mutation-agnostic, methylation-based predictor of clonal haematopoiesis burden that estimates predicted VAF (pVAF), to examine longitudinal associations between clonal burden and brain structure in the Lothian Birth Cohort 1936. In addition, to capture broader clonal burden, we developed CIMS, a novel methylation-based score derived from clonal haematopoiesis of indeterminate potential (CHIP) EWAS summary statistics, which reflects the epigenetic profile of both mutant and wild-type blood immune cells. Blood DNA methylation and structural MRI data were available at three timepoints spanning ages 73–79 years. The analytic dataset comprised 1,091 participants, of whom 808 (pVAF) and 836 (CIMS) contributed clonality data and 744 had brain MRI; 479–481 had complete baseline and longitudinal imaging measures with covariates. Trajectories were modelled with latent growth curve models fitted by full information maximum likelihood.

**Findings:** Longitudinal trajectories of clonality and grey matter volume were significantly associated over 6 years (Std. est. = −0.176, *p*_adj_ = 0.030), revealing a progressive, left-lateralised cortical atrophy pattern affecting medial temporal and parietal regions. This pattern showed significant spatial correspondence with established AD signatures (*r* = 0.357, *p* = 0.003), suggesting that clonality is linked to AD-related patterns of neurodegeneration. Subcortical analyses revealed nominal relative preservation of the caudate and did not detect a statistically significant association with hippocampal volume, suggesting potential divergence from the canonical AD subcortical profile. The CIMS confirmed global atrophy but revealed divergent tissue-type and regional patterns, suggesting the two measures may capture distinct aspects of CH biology.

**Interpretation:** Together, these findings suggest that CH burden may mark biological processes associated with structural brain atrophy in cognitively healthy older adults, with clonality showing a regional pattern that resembles the one reported in AD. CH burden may therefore be a candidate marker of adverse brain ageing; whether it tracks AD pathology in particular, rather than age-related neurodegeneration more broadly, remains to be tested.

## Introduction

Clonal haematopoiesis (CH) arises when haematopoietic stem cells acquire somatic mutations that confer a competitive advantage on the mutant cell relative to its neighbours, leading to progressive expansion of that clone within the blood. When clonal expansion of a haematopoietic driver gene variant reaches a variant allele frequency (VAF) of at least 2% in the absence of cytopenia or morphological abnormality, it is classified as clonal haematopoiesis of indeterminate potential (CHIP).^1^ Prevalence rises steeply with age: applying this threshold to whole-exome sequencing data, CH is detectable in approximately 6% of people aged 60–69, 10% of those aged 70– 79, and 18% of those aged 90 and over.^2,3^ The mutations involved arise most commonly in epigenetic regulators (*DNMT3A, TET2, ASXL1*), DNA damage response genes, or signalling pathway components. CHIP is an established risk factor for cardiovascular disease, haematologic malignancies, and all-cause mortality.^4^ The mechanisms linking CH to adverse outcomes are not fully understood, though dysregulated inflammatory signalling in CH-mutant myeloid cells has been proposed as a key contributor.^5,6^ These myeloid cells include monocyte-derived tissue macrophages such as infiltrating brain microglia, positioning CH as a possible systemic driver of age-related pathology.

The relationship between CH and neurodegeneration remains contentious. Some studies reported that CHIP conferred protection against cognitive impairment,^7,8^ while others identified CHIP as a risk factor for stroke and Parkinson’s disease,^9–14^ though exceptions exist.^15^ For Alzheimer’s disease (AD) specifically, studies have reported protective,^16–18^ null,^15,19,20^ and detrimental associations.^21–23^ Notably, distinct analyses of the same cohort, the Alzheimer’s Disease Sequencing Project (ADSP), have yielded opposite conclusions: Bouzid et al. reported that CHIP was protective against AD^16^ whereas others suggested increased AD risk.^21,22^ This heterogeneity likely reflects differences in participant selection, CH definitions and VAF cutoffs, mutation-specific biology, and clinical endpoints, most of which rely on diagnoses or cognitive scores with limited resolution of regional brain pathology or temporal dynamics.

Structural neuroimaging offers finer-grained intermediate phenotypes (cortical atrophy, white matter change) but results remain inconsistent^9,24–27^: *DNMT3A*-CHIP has been linked to lower white matter hyperintensity (WMH) volume, suggesting a protective vascular effect,^24^ whereas CHIP overall has been linked to brain infarcts but not atrophy or WMH volume.^25^ Only one study has examined longitudinal relationships between imaging and baseline CHIP, linking mutations to incident microbleeds and white matter lesions.^27^ Critically, this gap reflects a measurement limitation: most longitudinal imaging cohorts derive CH status from a single baseline blood draw, so any “longitudinal” analysis simply propagates a static measure forward.

A smaller number of cohorts diverge from this pattern by collecting repeated blood DNA methylation alongside repeated brain imaging. The Lothian Birth Cohort 1936 (LBC1936)^28,29^ is one such resource, and its methylation data can be leveraged to infer CH burden longitudinally using our previously developed method, COMET (Clonal Observation from METhylation).^30^ COMET is mutation-agnostic, detecting clonal expansion via CpG sites whose methylation shifts predictably as clones expand, regardless of the underlying genetic driver. This matters because the conventional CHIP definition undercounts clonal expansion along two independent axes. Requiring a canonical driver gene misses the ∼78% of clonal expansions in elderly individuals that, by whole-genome sequencing estimates, lack one.^31^ Additionally, requiring a 2% variant allele frequency misses smaller clones: error-corrected sequencing at a 1% threshold detects clonal haematopoiesis in 62% of individuals aged 80 and older.^32^ Validated against sequencing-based VAF estimates (R² = 0.80),^30^ COMET enables scalable application to cohorts with existing methylation data and, crucially, supports longitudinal tracking of clonal dynamics from repeated methylation measures.

Here, we leveraged the LBC1936 cohort, which offers a unique dual-longitudinal design: blood DNA methylation and structural brain MRI were assessed at three timepoints spanning ages 73–79 years. This enabled us to examine (1) whether baseline clonality burden is associated with baseline brain volume, and (2) whether the rate of change in clonality burden is associated with the rate of change in brain volume. We tested clonality burden against global, tissue-type, regional cortical, and subcortical brain volumes, examining whether effects follow normative ageing patterns or preferentially target neurodegeneration-vulnerable regions. Finally, we developed a complementary predictor, the CHIP-associated immune methylation score (CIMS), built from CHIP epigenome-wide association study (EWAS) summary statistics from Kirmani et al.,^33^ to capture systemic epigenetic remodelling beyond clone size alone.

## Results

### Clonality burden of clonal haematopoiesis is associated with global brain and grey matter atrophy

The overall study design and analytical pipeline are summarised in Figure 1. Briefly, we leveraged the LBC1936, which provides repeated structural brain MRI and blood DNA methylation across three waves (ages 73, 76, and 79 years). Following preprocessing and quality control, the analytic dataset comprised 1,091 participants (Table S1); the pattern of missing data across waves and variables is shown in Figure S1. Structural MRI was available for 744 participants, of whom 658 had wave-2 baseline tissue volumes and 505 an estimable volumetric slope across waves 2–4. Brain volume metrics were derived from structural MRI as described in the Methods, with intercept and slope metrics representing the baseline value at wave 2 and the rate of volumetric change across waves 2–4, respectively (Figure S2). Clonality measurements were available for a subset of these: 808 participants contributed at least one pVAF measurement across waves 2–4, and 836 at least one CIMS measurement. COMET-predicted pVAF trajectories across the three waves are shown in Figure S3. Combining the two, 502 participants had both a wave-2 pVAF measurement and a wave-2 total brain volume, and 384 had both a wave-2 pVAF measurement and an estimable total brain volume slope (CIMS: 605 and 465); 479 (pVAF) and 481 (CIMS) had complete baseline and slope imaging data together with all base covariates. The brain–clonality estimates reported below therefore rest on this imaged subgroup rather than on the full dataset. Associations between brain volume trajectories and pVAF trajectories, at both the intercept–intercept level (baseline values) and slope–slope level (rates of change), were then examined using latent growth curve (LGC) modelling within a structural equation modelling (SEM) framework.

**Figure 1.**
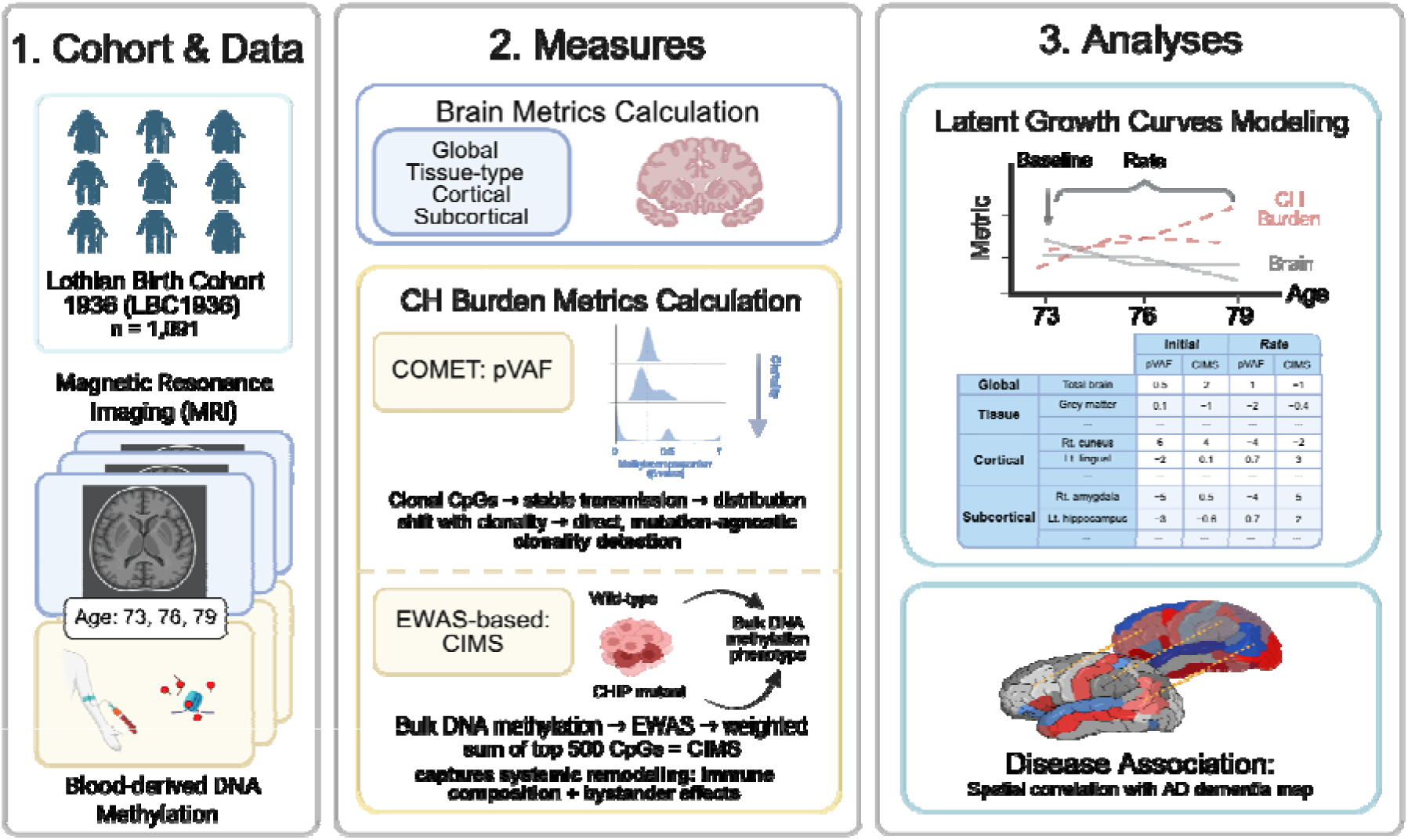
Schematic overview of the study design and analytical pipeline. Longitudinal data from the Lothian Birth Cohort 1936 (ages 73, 76, and 79 years) were integrated to assess the association between structural brain metrics and methylation-based clonal haematopoiesis (CH) burden, at both baseline level and rate of change. The workflow encompasses: Brain Metrics Calculation, comprising volumetric calculation of global, tissue-type, cortical, and subcortical brain metrics; CH Burden Metrics Calculation, including COMET-derived predicted variant allele frequency (pVAF), with clonality inferred from the β value distribution of selected CpGs, and the CHIP-associated immune methylation score (CIMS), calculated as a weighted sum of methylation β values at the top 500 CH-associated EWAS CpGs; and latent growth curve modelling, used to estimate baseline levels and rates of change for brain and CH burden metrics, and to test their association with each other and with spatial dementia maps (Disease Association). The analytic dataset comprised 1,091 participants, of whom 808 (pVAF) and 836 (CIMS) contributed at least one clonality measurement and 744 had at least one structural MRI measure (658 baseline volumes; 505 volumetric slopes). Complete baseline-and-slope imaging with covariates was available for 479 (pVAF) and 481 (CIMS) participants.

#### Baseline clonality burden is associated with lower baseline total brain volume

At the macrostructural level, total brain volume exhibits age-related decline,^34,35^ and is associated with cognitive deterioration in both physiological ageing of healthy brains^34,36,37^ and neurodegenerative conditions.^38^ We therefore investigated whether COMET-predicted VAF (pVAF), a clonality burden, correlated with global brain atrophy. Within this latent growth curve framework, the intercept–intercept association showed that pVAF was associated with lower total brain volume after adjusting for age, sex, and intracranial volume (ICV) (Std. est. = −0.270, *p*_adj_ = 0.023; Table 1), suggesting that greater clonal burden corresponds to reduced total brain volume at the baseline level. However, the slope–slope association between pVAF and total brain volume over the subsequent 6 years, after adjusting for age and sex, was not statistically significant (Std. est. = −0.084, *p* = 0.163; Table 1), indicating that clonal burden relates most clearly to baseline brain structure.

**Table 1.** Associations between clonal haematopoiesis features (pVAF and CIMS) and total brain volume at baseline level (intercept) and rate of change (slope), adjusted for age and sex. Models were fitted by full information maximum likelihood. Clonality data were contributed by 808 participants (pVAF) and 836 (CIMS); complete baseline and slope total brain volume data with covariates were available for 479 and 481 respectively. See Methods for the distinction between these counts and lavaan’s reported N. Unstandardised associations are shown (Estimate) and their corresponding standard error (SE) and *p*-value. Nominally significant associations (*p* < 0.05) are in bold typeface. CH = clonal haematopoiesis; pVAF = COMET-predicted variant allele frequency; CIMS = CHIP-associated immune methylation score; *p*_adj_ = Benjamini–Hochberg FDR-adjusted *p*-value.

| CH Feature | Growth Parameter | Estimate | SE | *p*-value | Std. est. | *P*<sub>adj</sub> |
| --- | --- | --- | --- | --- | --- | --- |
| pVAF | Intercept | **-0.249** | **0.105** | **0.018** | **-0.270** | **0.023** |
| pVAF | Slope | -0.444 | 0.318 | 0.163 | -0.084 | 0.163 |
| CIMS | Intercept | **-0.049** | **0.014** | **< 0.001** | **-0.385** | **0.003** |
| CIMS | Slope | **-0.145** | **0.048** | **0.003** | **-0.187** | **0.005** |

#### Grey matter shows accelerated atrophy while white matter measures show no association

Cortical grey and white matter exhibit distinct cellular compositions, metabolic profiles, and vulnerability to neurodegeneration.^39^ In addition, their volumes are independently associated with cognitive function.^34,40^ To capture compartment-specific effects that may be obscured in whole-brain analyses, we separately examined grey and white matter volumes. This revealed divergent patterns across compartments (Table 2, Figure S4). While the intercept– intercept association between grey matter (GM) volume and pVAF showed only a trend (Std. est. = −0.128, *p* = 0.088), the slope–slope association was significant, with clonality burden tracking the rate of GM volume change over 6 years (Std. est. = −0.176, *p*_adj_ = 0.030). This longitudinal specificity suggests that clonality burden, at the GM level, may be more strongly associated with continuing neurodegenerative processes than with static structural differences. In contrast, we found no significant intercept–intercept or slope–slope associations between clonality burden and white matter measures, including normal-appearing white matter (NAWM) volume and white matter hyperintensity (WMH) volume, a cerebral small vessel disease marker seen in vascular dementia.^41^

**Table 2.** Associations between clonal haematopoiesis features (pVAF and CIMS) and brain tissue-type measures at baseline level (intercept) and rate of change (slope), adjusted for age and sex. Models were fitted by full information maximum likelihood. Clonality data were contributed by 808 participants (pVAF) and 836 (CIMS); complete baseline and slope tissue-type volume data with covariates were available for 479 and 481 respectively. See Methods for the distinction between these counts and lavaan’s reported N. Unstandardised associations are shown (Estimate) and their corresponding standard error (SE) and *p*-value. Standardised estimates allow direct effect size comparisons (Std. est.). Nominally significant associations (*p* < 0.05) are in bold typeface. CH = clonal haematopoiesis; pVAF = COMET-predicted variant allele frequency; CIMS = CHIP-associated immune methylation score; GM = grey matter volume; NAWM = normal-appearing white matter volume; WMH = white matter hyperintensity volume. *p*_adj_ = Benjamini–Hochberg FDR-adjusted *p*-value.

| CH Feature | Growth Parameter | Tissue type | Estimate | SE | *p*-value | Std. est. | *P*<sub>adj</sub> |
| --- | --- | --- | --- | --- | --- | --- | --- |
| pVAF | Intercept | GM | -0.235 | 0.138 | 0.088 | -0.128 | 0.264 |
| pVAF | Intercept | NAWM | -0.005 | 0.146 | 0.975 | -0.003 | 0.975 |
| pVAF | Intercept | WMH | -0.002 | 0.002 | 0.295 | -0.048 | 0.442 |
| **pVAF** | **Slope** | **GM** | **-0.958** | **0.341** | **0.005** | **-0.176** | **0.030** |
| pVAF | Slope | NAWM | -0.150 | 0.136 | 0.273 | -0.065 | 0.442 |
| pVAF | Slope | WMH | -0.696 | 0.735 | 0.344 | -0.053 | 0.459 |
| **CIMS** | **Intercept** | **GM** | **-0.093** | **0.018** | **< 0.001** | **-0.367** | **< 0.001** |
| CIMS | Intercept | NAWM | 0.015 | 0.020 | 0.453 | 0.067 | 0.543 |
| CIMS | Intercept | WMH | 0.0003 | 0.0003 | 0.205 | 0.057 | 0.410 |
| CIMS | Slope | GM | -0.066 | 0.052 | 0.203 | -0.082 | 0.410 |
| **CIMS** | **Slope** | **NAWM** | **-0.048** | **0.021** | **0.024** | **-0.140** | **0.096** |
| CIMS | Slope | WMH | -0.037 | 0.113 | 0.741 | -0.019 | 0.809 |

### Clonal haematopoiesis is associated with a regional pattern of cortical atrophy that spatially overlaps with Alzheimer’s disease signatures

Having established that clonality burden is associated with accelerated atrophy in the grey matter compartment, we next investigated whether this tissue loss exhibits diffuse or region-specific patterns, as anatomical selectivity could reveal underlying pathogenic processes.^42,43^ To examine whether clonality burden shows regional specificity in its associations with brain structure, we conducted region-wise analyses of cortical volume across 68 anatomically defined regions using the Desikan–Killiany atlas (Figure 2A, Table S2).

**Figure 2.**
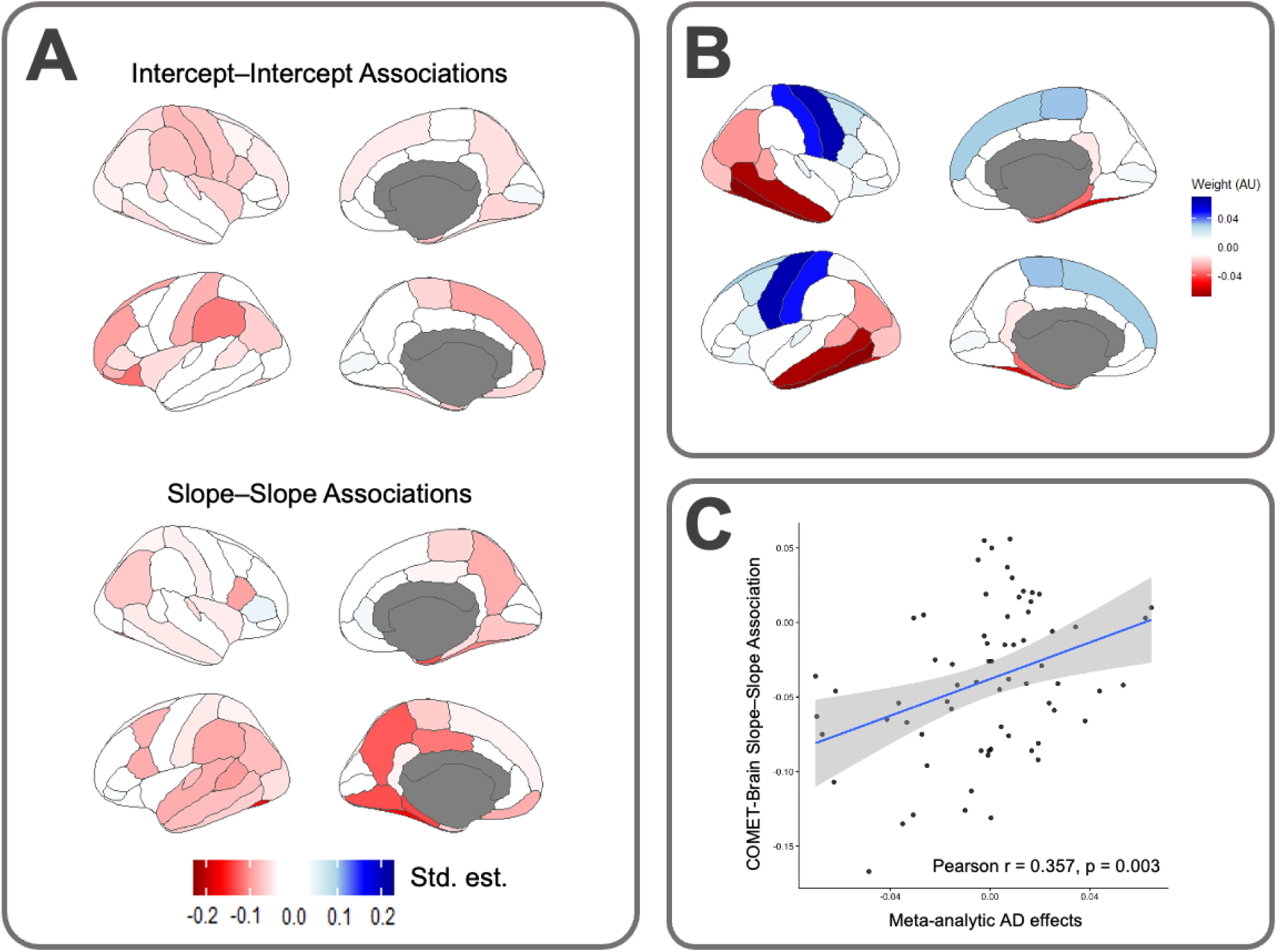
Regional cortical atrophy associated with COMET-predicted VAF recapitulates Alzheimer’s disease vulnerability patterns. (A) Latent growth curve modelling reveals region-specific associations between COMET-derived VAF and cortical volume across Desikan–Killiany (DK) parcellations. Upper panel: cross-sectional associations (intercept–intercept). Lower panel: longitudinal trajectory associations (slope–slope). Standardised estimates (Std. est.) are colour-coded: red indicates negative associations (higher VAF → accelerated volume loss), blue indicates positive associations. Grey regions denote areas excluded from analysis. (B) Meta-analytic cortical vulnerability map for AD dementia derived from case-control cortical surface area studies (*n* = 4,774).^44^ Effect sizes (arbitrary units) represent regional surface area reduction in AD (vulnerability); blue indicates relatively spared or larger surface area (resilience). (C) Region-wise correlation between pVAF-associated longitudinal atrophy (slope–slope association estimate from panel A, lower) and meta-analytic AD vulnerability (panel B). Each point represents one DK region (*n* = 68 parcels). The positive correlation (Pearson *r* = 0.357, *p* = 0.003) demonstrates that cortical regions most vulnerable to clonality-related atrophy overlap significantly with canonical AD-vulnerable regions. Blue line: linear fit; grey ribbon: 95% confidence interval.

#### Associations at baseline level revealed focal clusters in which higher predicted VAF was associated with lower cortical volume

In the left hemisphere (Figure 2A), the effects were nominally significant in the lateral orbitofrontal cortex and supramarginal gyrus (Figure S5), with additional effects in the superior frontal, rostral middle frontal, pars orbitalis, and postcentral regions. In the right hemisphere (Figure 2A), prominent effects were seen in postcentral cortex, pars opercularis, precentral gyrus, supramarginal gyrus, and insula. Overall, at baseline level, there was comparable involvement of both hemispheres.

#### Association of trajectories reveals a left-lateralised medial temporal and parietal signature

Given our global brain analysis finding that clonality burden is robustly associated with progressive GM loss, we hypothesised that the biologically relevant anatomical signatures would emerge in the longitudinal data. Accordingly, we identified regions where predicted VAF (pVAF) was associated with accelerated atrophy over time. Longitudinal associations between clonality burden and cortical change showed a qualitatively strong left-lateralised pattern of accelerated atrophy that was distinct from the baseline profile (Figure 2A). The most pronounced effects were observed in the left hemisphere, particularly in the fusiform, lingual, and parahippocampal gyri, and the precuneus. Right hemisphere effects were largely confined to the entorhinal cortex and fusiform gyrus. Additional bilateral effects were noted in the pars opercularis and the inferior parietal lobule.

#### CH-associated atrophy spatially overlaps with Alzheimer’s disease signatures

The longitudinal atrophy pattern identified above, preferentially affecting medial temporal and parietal regions, resembles cortical signatures reported in AD. To quantitatively determine whether this pattern mirrors known neurodegenerative signatures, we calculated the spatial correlation between the COMET-associated longitudinal cortical volume map and a meta-analytic map of regional case-control AD surface area effects.^44^ In the slope–slope association, we identified a significant spatial correlation (*r* = 0.357, *p* = 0.003; Figure 2A [lower panel], 2B and 2C). In contrast, the correlation against the intercept–intercept association did not reach statistical significance (*r* = −0.165, *p* = 0.178). This effect size (*r* = 0.357) is comparable in magnitude to the spatial correlation reported between family history of AD and clinical AD atrophy patterns (*r* = 0.33),^45^ a relationship attributable to genetic risk. The similarity in effect size suggests that the topographic overlap between CH-associated atrophy and AD atrophy is not trivial and falls within a range considered biologically meaningful in other AD-relevant contexts. Collectively, we characterise clonality burden as being associated with a distinct, progressive pattern of brain atrophy that spatially overlaps with Alzheimer’s disease pathology, distinguishing it from generalised ageing patterns.

### Subcortical preservation distinguishes clonal burden from Alzheimer’s pathology

Building on the cortical AD-like signature identified above and given that specific subcortical structures are hallmark sites of pathology in neurodegenerative disorders (most notably the hippocampus in AD and the striatum in Parkinson’s disease), we next sought to determine whether CH-associated structural changes extended to these subcortical grey matter volumes. Using the same longitudinal modelling framework applied to the cortical regions of interest (ROIs), we examined associations across the bilateral accumbens, amygdala, caudate, hippocampus, pallidum, putamen, thalamus, and ventral diencephalon. We observed a small number of focal subcortical associations. At baseline level, higher clonality burden was associated with larger left nucleus accumbens volume (Table S3; Std. est. = 0.114, *p* = 0.008, *p*_adj_ = 0.208). Longitudinally, higher clonality burden was associated with more positive caudate volume slopes in both hemispheres (Left caudate: Std. est. = 0.155, *p* = 0.013, *p*_adj_ = 0.208; Right caudate: Std. est. = 0.128, *p* = 0.040, *p*_adj_ = 0.427), indicating relative preservation (i.e., less decline) of caudate volume over follow-up among individuals with higher clonality burden. No other subcortical region showed evidence of association at the nominal significance threshold (Table S3).

In summary, the lack of detectable hippocampal associations and the nominal preservation of the caudate suggest that while the cortical signature of pVAF correlates with AD-vulnerable spatial patterns, it does not fully recapitulate the subcortical profile characteristic of clinical AD in this cohort.

### A complementary EWAS-based predictor confirms global atrophy but reveals divergent regional trajectories

#### A complementary EWAS-based predictor captures systemic epigenetic remodelling beyond clonal size

While COMET directly quantifies clone size, it does not capture the broader systemic epigenomic remodelling triggered by CHIP mutations across the entire blood compartment. To assess this complementary signal, we constructed another predictor using CHIP EWAS summary statistics from Kirmani et al.^33^ Specifically, we computed a weighted methylation score as the sum of methylation β values at the top 500 CHIP-associated CpGs multiplied by their respective effect sizes, resulting in the CHIP-associated immune methylation score (CIMS). CIMS may reflect broader CHIP-associated blood methylation remodelling, potentially including changes in immune-cell composition, inflammatory state, and methylation changes in non-mutant bystander cells,^46,47^ although these components cannot be separated in bulk DNAm data.

CIMS was consistently distributed relative to COMET-predicted VAF across waves (Figure S6, left) and showed moderate-to-strong within-wave correlations (Figure S6, right), indicating that the two measures capture overlapping but not identical aspects of CH burden.

#### Both CH features, pVAF and CIMS, converge on global atrophy but diverge regionally

Global brain associations with CIMS were qualitatively similar to COMET-predicted VAF (pVAF) (Table 1), showing significant baseline associations with total brain volume (Std. est. = −0.385, *p*_adj_ = 0.003) and, in contrast to COMET-derived pVAF, significant longitudinal association with brain atrophy (Std. est. = −0.187, *p*_adj_ = 0.005). Further grey/white matter analyses (Table 2, Figure 3A) showed that for grey matter volume, CIMS was associated at baseline (Std. est. = −0.367, *p*_adj_ < 0.001) but not longitudinally (Std. est. = −0.082, *p* = 0.203), contrasting with the pVAF pattern of strong longitudinal but weaker baseline effects (Figure S7). Notably, the EWAS-based score, CIMS, was nominally associated with longitudinal atrophy of normal-appearing white matter (NAWM; Std. est. = −0.140, *p* = 0.024, *p*_adj_ = 0.096), a pattern that is also associated with cognitive decline.^40^ Regional analyses revealed divergent spatial patterns compared with pVAF (Figure 3B; Table S4). At baseline, CIMS was broadly distributed across frontal (particularly rostral middle frontal and lateral orbitofrontal cortices), temporal, and parietal regions. Longitudinally, CIMS was associated with paradoxical positive volume changes in sensorimotor and temporal regions, opposite to the accelerated atrophy pattern observed with COMET. We found no spatial correlation between CIMS and the meta-analytic AD dementia map^44^ (intercept–intercept: *r* = −0.089, *p* = 0.469; slope–slope: *r* = −0.143, *p* = 0.243). Subcortical associations were confined to baseline effects in the left accumbens (Intercept: Std. est. = 0.103, *p* = 0.016, *p*_adj_ = 0.512), with no nominally significant longitudinal subcortical slope associations (Table S5).

**Figure 3.**
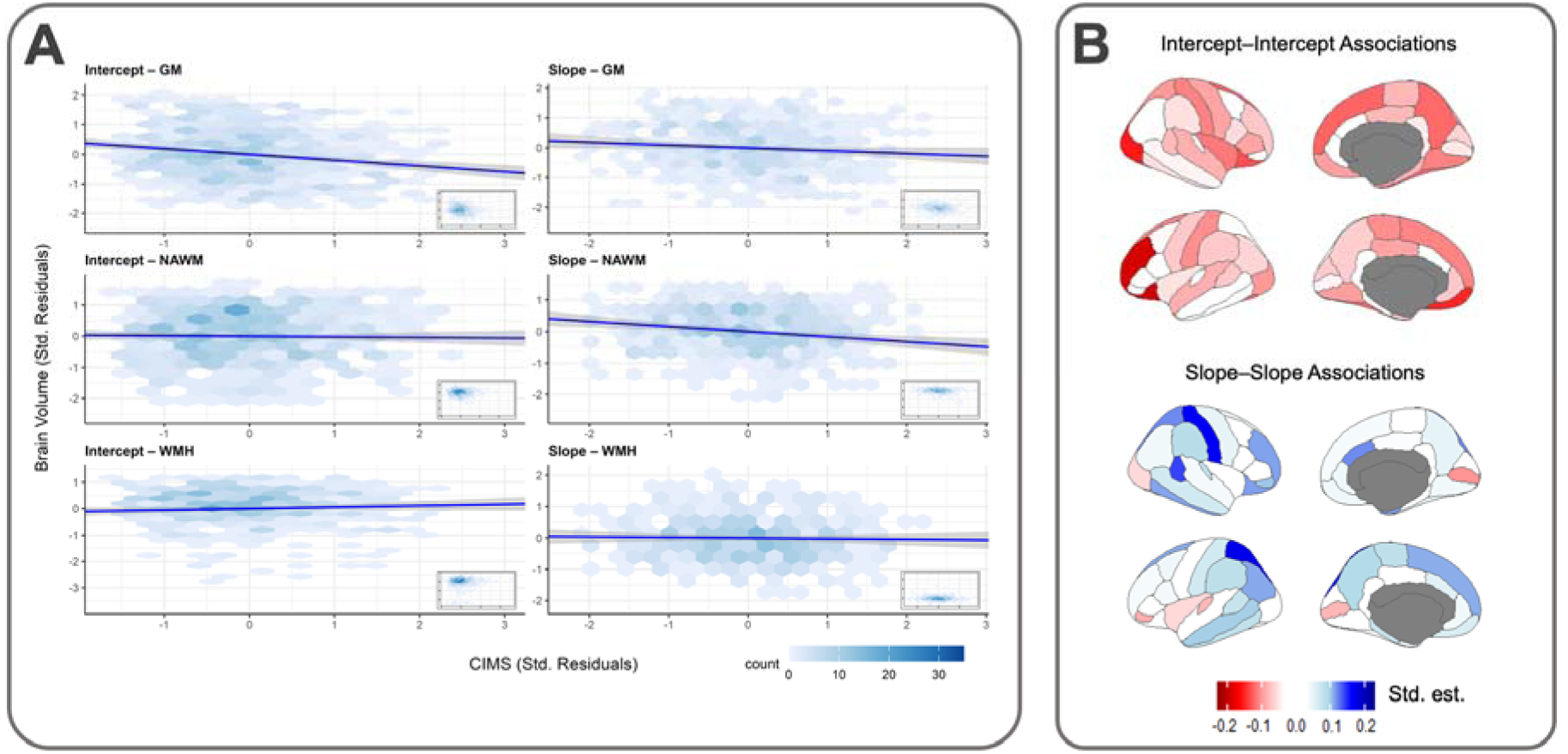
Associations between CHIP-associated immune methylation score and brain volumetry: baseline and rate of change. (A) Scatter plots show associations between CIMS and tissue volumes (WMH, GM, NAWM) for baseline levels (intercept) and rates of change (slope) from latent growth curve models. Both variables were adjusted for age, sex; axes show standardised residuals. For visualisation, each panel’s axes are clipped to the central 95% (2.5th–97.5th percentile) of its data with a small margin added; regression lines and 95% CIs are fit using the complete, unclipped data. Hex bins indicate point density on a shared count scale across all panels; inset panels (bottom-right of each plot) show the corresponding unclipped hex-binned distribution of all data points, without a fitted line. (B) Region-specific associations between CIMS and cortical volumes across Desikan–Killiany parcellations. Top: baseline (intercept–intercept); bottom: longitudinal (slope–slope). Red = negative association (higher score → lower baseline volume or faster rate of atrophy); blue = positive association. Grey = excluded regions. CIMS = CHIP-associated immune methylation score.

In summary, while both measures converge on global atrophy, their divergent regional signatures suggest that clonal expansion and pan-blood epigenetic remodelling represent distinct biological pathways.

### Sensitivity analysis: robust associations after adjustment for blood cell-type counts and technical batch

As both predictors utilise information from bulk methylation data, the dynamics of cell-type counts and technical variations could theoretically contribute to deviations in predicted values to varying extents, depending on the context and assumptions. Therefore, we conducted sensitivity analyses by regressing out leucocyte counts alongside technical batch effects (Table S6 and S7). For global atrophy, the direction of associations remained similar for both COMET-predicted VAF and the CHIP-associated immune methylation score (CIMS). For grey/white matter atrophy, overall results were similar, except that longitudinal NAWM associations with CIMS were no longer significant after the adjustment. At the regional cortical level, the adjustment attenuated the effect sizes. For pVAF, the four left-hemisphere regions identified above (fusiform, lingual, parahippocampal, and precuneus) fell below nominal significance (Table S8). For CIMS, two baseline regional associations, namely left rostral middle frontal and right lateral occipital, lost significance under FDR correction (Table S9). The longitudinal spatial correspondence with the meta-analytic AD map, for pVAF, was attenuated and no longer reached significance (*r* = 0.230, *p* = 0.059), consistent with the attenuation observed at the regional level (Table S8). Interestingly, the baseline pattern showed a weak inverse correspondence (*r* = −0.277, *p* = 0.022), which we do not interpret further given that the unadjusted baseline correlation was null (*r* = −0.165, *p* = 0.178) and no baseline region survived correction. For CIMS, there was no spatial correspondence under adjustment either at baseline or longitudinally.

In summary, using COMET, our investigation in the LBC1936 demonstrates that clonality burden from clonal haematopoiesis is significantly associated with global and grey matter brain atrophy. The longitudinal cortical regional association map is spatially correlated with the AD pathology map, whereas subcortical analysis fails to capture a similar correlation. The EWAS-based predictor captures a shared signal of global macrostructural decline but shows divergent regional trajectories, implying that different aspects of CH are being captured. Lastly, although theoretically influenced by blood cell-type counts, the two predictors retain their global and spatial properties after cell-type adjustment, albeit with some attenuation of focal regional effect sizes.

## Discussion

In this study, we used methylation-based clonal haematopoiesis (CH) predictors to examine associations between clonal haematopoiesis and macrostructural brain changes assessed through longitudinal MRI in the LBC1936. The LBC1936 represents a generally cognitively healthy ageing cohort with prevalence rates of mild cognitive impairment (MCI) of approximately 15% at wave 3 (age 76) and 17% at wave 4 (age 79),^48^ while dementia prevalence in the age group 75–79 years was approximately 4.5%,^49^ meaning our observed associations between clonality burden and brain atrophy occur primarily in the context of otherwise normal cognitive ageing rather than in participants affected by overt neurodegeneration.

Our findings contribute to an emerging but complex literature on CH and brain structure. Previous studies in community-dwelling populations have reported seemingly contradictory results. Lee et al. found that clonal haematopoiesis with *DNMT3A* mutations was associated with lower WMH volume, which they interpreted as protective because WMHs typically reflect chronic hypoperfusion or microvascular damage.^24^ In contrast, Li et al.^25^ reported that CHIP was associated with large brain infarcts in a community-based cohort of 1,229 participants (18.2% with CHIP), but found no association with WMH volume or brain atrophy. The divergent findings from Lee et al. and Li et al., in which *DNMT3A* CHIP showed protective effects against WMH yet CHIP overall was associated with large infarcts, underscore the complexity of CH effects on brain-related pathology and suggest that specific mutations or clonal characteristics may yield distinct patterns of cerebrovascular versus neurodegenerative pathology.

Our study differs from these previous investigations in several key ways. First, we employed longitudinal repeated MRI assessments rather than single cross-sectional measurements, allowing us to capture progressive brain changes over time rather than static structural differences. Second, we utilised COMET (Clonal Observation from METhylation), a methylation-based predictor that measures clonality burden continuously and captures clonal expansion regardless of the underlying cause. Because COMET operates in a mutation-agnostic manner, it effectively identifies CH cases that might be missed by traditional sequencing, providing more comprehensive detection coverage. This is particularly important given that whole-genome sequencing studies demonstrate that approximately 78% of clonal expansions in elderly individuals lack mutations in known driver genes,^31^ which suggests that current targeted sequencing approaches miss substantial clonal heterogeneity. The dual-longitudinal design and the mutation-agnostic approach together increase the granularity of our findings relative to previous studies.

Longitudinally, the regional patterns associated with the COMET predictor partially overlap with the established topography of age-related cortical atrophy, particularly in lateral temporal regions.^50^ This suggests that CH burden is associated with a pattern qualitatively resembling accelerated normative ageing. Furthermore, we observed a significant correlation between COMET and the meta-analytic AD dementia map, indicating that the longitudinal COMET–cortex pattern significantly overlaps with AD-related atrophy in regions including the parahippocampus and fusiform gyrus.^43,44^ Of note, the AD-like topography is directionally consistent but not robust to cell-composition and technical batch adjustment, so it should be treated as preliminary.

Surprisingly, subcortical analysis did not show a significant association in AD-vulnerable areas such as the hippocampus. This divergence from our cortical-level results implies that the association does not fully recapitulate AD pathology. Together, these results indicate that clonality burden is associated with both a pattern resembling accelerated normative ageing and cortical changes overlapping those characteristic of AD.

The paradoxical preservation of caudate volume in individuals with higher clonality burden is intriguing. While speculative, this could reflect clonality-associated alterations in dopaminergic signalling or microglial function that differentially affect striatal versus cortical circuits. Alternatively, this may reflect survivor bias if individuals with both high clonality burden and striatal pathology were more likely to develop Parkinson’s-related syndromes and were excluded from this cognitively healthy cohort. Of note, as the test did not survive correction for multiple comparisons, the result should be regarded as provisional.

We developed a complementary methylation-based score, the CHIP-associated immune methylation score (CIMS), from the clonal haematopoiesis of indeterminate potential (CHIP) EWAS summary statistics,^33^ allowing us to compare clone-size-based and broader epigenomic-remodelling-based signals within the same cohort; this is, to our knowledge, the first study to triangulate CH-associated neurodegeneration using two complementary, non-redundant methylation-derived instruments. While COMET and CIMS diverged in their regional trajectories, with CIMS showing a lack of AD-spatial correlation, they converged robustly on global brain atrophy at baseline. This convergence across two distinct methylation-based methodologies reinforces the fundamental link between haematopoietic health and macrostructural brain integrity. The regional divergence suggests that while ‘clonal’ expansion (pVAF) was associated with AD-like cortical volume loss, the broader ‘episignature’ of the blood might reflect different systemic processes, such as white matter changes, which we observed nominally in CIMS–NAWM longitudinal associations.

A major strength of this study is the availability of longitudinal cohort data with repeated brain imaging assessments spanning 6 years across three timepoints, allowing us to distinguish cross-sectional associations from progressive changes over time.

However, certain limitations should be acknowledged. First, our methylation-based CH predictors lack mutation-specific resolution. As previous studies have demonstrated that different CH-driving mutations can have different, or even opposite, effects on clinical outcomes, mutation-specific information would enable us to identify which particular mutations are associated with specific brain atrophy patterns. The continuous, pan-CH nature of COMET provides comprehensive burden assessment but sacrifices this mutation- and genotype-specific resolution. Second, only 744 of 1,091 participants had structural MRI and 479–481 had complete baseline and slope imaging with covariates, so the brain associations rest on a subgroup that may not be representative of the full cohort. Lastly, while the spatial correlation with the AD-dementia map is notable, spatial overlap does not inherently imply shared molecular aetiology such as amyloid or tau pathology.

Future research should validate these findings in independent cohorts using the same methylation-based methodology (particularly COMET) to establish generalisability across populations and age ranges. While our analyses examined individual brain regions separately, complementary approaches using multivariate pattern analysis, such as latent variable models^43,50^ that identify coordinated atrophy across distributed networks, could reveal whether CH preferentially targets specific brain systems rather than isolated structures. Additionally, examining CH associations with other neuroimaging modalities, including functional MRI and connectivity measures, could provide deeper mechanistic insights into how clonal expansion influences neural network integrity and function. PET imaging or cerebrospinal fluid (CSF) biomarkers are needed to determine whether clonality burden directly correlates with traditional AD hallmarks. Integration of mutation-specific information with neuroimaging would allow investigation of whether particular CH-driving mutations show regional selectivity in their neurological effects.

## Conclusion

This study shows that methylation-based inference of clonal burden is a workable route to studying clonal haematopoiesis where serial sequencing is unavailable. COMET, developed in our laboratory, is mutation-agnostic. CIMS, built independently from published CHIP EWAS statistics, captures a broader blood episignature. The two are related but not interchangeable. In LBC1936, greater clonal burden was associated with lower total brain volume at age 73 and accelerated grey matter loss over follow-up. Exploratory regional analyses suggested a left-lateralised medial temporal and parietal gradient partially overlapping with Alzheimer’s disease atrophy. Replication in independent cohorts, integration with mutation-level sequencing, and pairing with amyloid/tau biomarkers will determine whether clonal burden marks AD pathology or a parallel route to cortical vulnerability.

## Methods

### Cohort and Participants

The Lothian Birth Cohort 1936 (LBC1936) is a prospective longitudinal study of individuals born in 1936 and residing in the Edinburgh area, who underwent repeated waves of clinical examination, blood collection, and structural brain MRI. The present analysis used waves 2–4, corresponding to mean ages of approximately 73, 76, and 79 years. Longitudinal blood-derived DNA methylation (DNAm) spans all three waves; applying COMET to these profiles enables longitudinal characterisation of clonal burden.

### COMET-derived predicted VAF

To estimate the variant allele frequency (VAF) from bulk DNA methylation data, we applied the COMET algorithm as previously described by Crofts et al.^30^ Briefly, for a predefined set of CpG sites with a known population mean methylation proportion, *p*, the algorithm assumes the underlying allele states in the population follow Hardy–Weinberg Equilibrium (HWE). The expected proportions of fully methylated (*P*_MM_), mixed (*P*_MU_), and fully unmethylated (*P*_UU_) cells are defined as *p*^2^*, 2p(1−p)*, and *(1−p)*^2^, respectively.

For a given individual *i*, the measured methylation proportion *x_i,j_* at CpG site *j* is assigned to an expected allele state based on HWE proportions. Site-specific clonality estimates, *v_i,j_*, are calculated based on the deviation of *x_i,j_* from the population mean:

For sites assigned to the *MM* state:

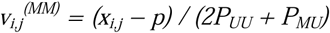

For sites assigned to the *UU* state:

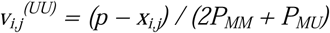

Estimates assigned to the *MU* state are excluded due to weaker signal-to-noise ratios. A raw clonality score, *C_i_*, is then calculated for the individual by averaging the *v_i,j_^(MM)^* estimates (if *p* < 0.5) or the *v_i,j_^(UU)^*estimates (if *p* > 0.5). Prior to averaging, the highest 10% of site-specific estimates are trimmed to mitigate the influence of extreme outliers.

Finally, to calibrate the raw score to targeted sequencing-derived clonal proportions, a linear adjustment is applied:

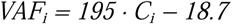

### MRI Acquisition and Processing

Lothian Birth Cohort 1936 participants underwent MRI scanning at the same clinic using the same scanner: a GE HealthCare Signa LX 1.5 T Horizon HDx clinical scanner equipped with a manufacturer-supplied 8-channel phased-array head coil. Further details on the structural imaging protocol for the LBC1936 cohort are provided in Wardlaw et al.^51^ T1-weighted images (3D IR-Prep FSPGR) were acquired in 160 coronal slices with a 256 mm field of view and a matrix size of 192 × 192 pixels (zero-filled to 256 × 256), resulting in a spatial resolution of 1 × 1 × 1.3 mm. Imaging parameters included a repetition time of 10 ms, echo time of 4 ms, and inversion time of 500 ms.

Regional cortical and subcortical volumetric segmentation was performed using FreeSurfer v5.3 (http://surfer.nmr.mgh.harvard.edu/), yielding 34 left/right pairs of cortical regions of interest (ROIs) (68 ROIs total, Desikan–Killiany atlas^52^) and subcortical structures. Brain volumetric measures, including total brain volume (TBV), grey matter (GM), white matter hyperintensity (WMH), and normal-appearing white matter (NAWM) volumes, were quantified using a semi-automated multispectral image processing approach.^53^

### Association Analyses

To investigate the relationship between DNAm-based predictors of clonal haematopoiesis (CH) and brain ageing, we employed latent growth curve modelling (LGCM) within a structural equation modelling (SEM) framework. Specifically, we modelled the association between COMET-predicted VAF (or CIMS) and the brain volumes calculated as previously described. The SEM framework consisted of two components: a measurement model and a structural regression model. While the measurement model remained consistent across all analyses, the structural regression models were tailored to the specific brain volumes under investigation.

### Measurement Model

The longitudinal trajectory of the DNAm-predicted variant allele frequency (VAF) was defined by two latent factors: an intercept (*VAF_I_*), representing the baseline VAF level at wave 2, and a slope (*VAF_S_*), representing the rate of change in VAF across waves 2, 3, and 4. The measurement model for the DNAm VAF at time *t* for individual *i* was defined as:

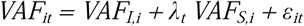

The factor loadings (λ_t_) were fixed to 0, 3.69, and 6.76 to reflect the mean years of follow-up between the respective waves.

### Structural Regression Model

We tested the associations between the latent VAF trajectories and brain structural measures. For global brain analyses, we regressed *VAF_I_*and *VAF_S_* onto baseline total brain volume (*TBV_I_*) and its longitudinal slope (*TBV_S_*). The structural equations were defined as follows:

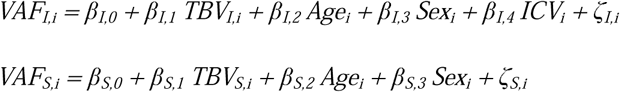

The coefficients β_I,1_and β_S,1_ estimate the associations between the VAF and brain volume latent factors. Because both variables are modelled as intercept (baseline level) and slope (rate of change) factors, we refer to β_I,1_throughout as the intercept–intercept association (the relationship between baseline level of VAF and the baseline level of brain volume at wave 2) and β_S,1_ as the slope–slope association (the relationship between the rate of change in VAF and the rate of change in brain volume).

For tissue-type level measurements, including grey matter (GM), normal-appearing white matter (NAWM), and white matter hyperintensities (WMH), the measurement model construct remained identical. However, the structural regression models incorporated all tissue types simultaneously as covariates:

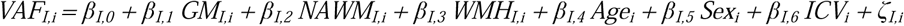

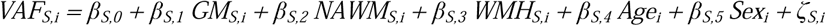

To examine regional specificity, we fitted separate SEMs for 68 cortical regions (defined by the Desikan– Killiany atlas) and various subcortical structures. We extracted standardised estimates (Std. est.) of beta weights (β) and corresponding p-values to evaluate the associations between regional volume (*V^r^*) intercepts/slopes and VAF trajectories. The regional (*r*) models were formulated as follows:

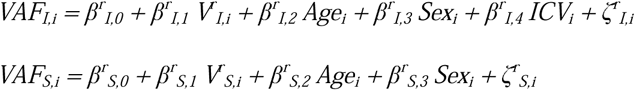

### Model Fitting

All models were fitted using the *growth()* function from the *lavaan* package^54^ in R. Default parameters were utilised, with the exception of the missing data handling parameter, which was set to missing = “ml.x” to accommodate missing data via Full Information Maximum Likelihood under a Missing At Random (MAR) assumption. Under this setting lavaan does not delete cases with missing exogenous covariates; the covariates instead enter the likelihood with freely estimated means and covariances, and each participant’s likelihood is evaluated from the variables observed for them. Because sex was recorded for every participant, no case met lavaan’s deletion criterion and the software therefore reported N = 1,091 for all models. This figure should not be read as the number of participants informing the reported associations: 283 (pVAF) and 255 (CIMS) participants had no clonality measurement at any wave and contributed only to the estimated covariate means and covariances, and participants contribute information solely to parameters involving variables they have. We therefore report throughout the number contributing clonality data (808 for pVAF, 836 for CIMS). Refitting the primary models in these restricted samples changed no standardised estimate by more than 0.005. Model fit was evaluated using Comparative Fit Index (CFI), Tucker–Lewis Index (TLI), and Root Mean Square Error of Approximation (RMSEA). Fit statistics across regional cortical models are provided in Figure S8.

### Sensitivity Analysis: Cell-Type Counts and Technical Batch Correction

To ensure that the observed associations were not driven by changes in blood cell composition, we conducted sensitivity analyses by correcting the DNAm predictors for major immune cell counts. Absolute counts of neutrophils, lymphocytes, monocytes, eosinophils, and basophils were obtained using standard automated clinical haematology analysers.

Corrected DNAm VAF values were derived by extracting the residuals from a linear regression model in which the raw VAF was the dependent variable and immune cell counts alongside technical batch effects were the independent variables.

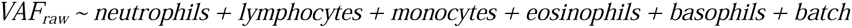

The primary association analyses (global, tissue-type, regional cortex, and subcortex) were subsequently repeated using these cell-corrected residuals to verify the robustness of the findings.

### Spatial Correlation with External Dementia Brain Map

To determine whether the spatial pattern of CH-related brain atrophy mirrors patterns characteristic of neurodegenerative disease, we performed a spatial correlation analysis using an external meta-analytic dementia map derived from a subset of cohorts included in a published Alzheimer’s disease (AD) case-control meta-analysis of regional cortical surface area.^44^ Of the 11 cohorts included in the original meta-analysis, five were used in the present study: the Alzheimer’s Disease Neuroimaging Initiative (ADNI; split into ADNI1 and ADNI2+GO+3 based on MRI scanner strength^55,56^), the Australian Imaging, Biomarkers and Lifestyle study,^57^ the Alzheimer’s Disease Repository Without Borders (ARWIBO^58,59^), and the Open Access Series of Imaging Studies 3 (OASIS-3^60^), totalling *n* = 4,774 participants (*n* = 790 AD; *n* = 3,984 controls). Pearson correlation coefficients were calculated between two sets of region-specific values across 68 cortical regions: (1) standardised estimates (Std. est.) of beta weights (β) derived from our latent growth curve models, reflecting the effect of clonality burden on the intercept or slope of cortical volume change; and (2) meta-analytic beta weights extracted from the dementia map.

### CHIP-associated Immune Methylation Score (CIMS)

To compute CIMS, we implemented a scoring approach based on a previously published multi-cohort EWAS of CHIP (*n* = 8,196).^33^ LBC1936 was not among the contributing cohorts, ensuring no sample overlap between discovery and validation datasets.

Briefly, the original EWAS identified CpG sites whose methylation levels were significantly associated with binary CHIP status, defined by a variant allele frequency (VAF) threshold of ≥2% in driver mutations. From the published EWAS results, the top 500 CHIP-associated CpGs were selected based on statistical significance of their association with CHIP status.

The CIMS for each individual was computed as a weighted linear sum:

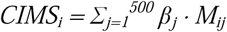

where β_j_ = the EWAS regression coefficient for CpG *j*, representing the mean methylation difference between CHIP-positive and CHIP-negative individuals; *M*_ij_ = the observed DNA methylation value (beta value) at CpG *j* for individual *i*; higher scores reflect a methylation profile more consistent with CHIP-positive status.

All association analyses, model fitting procedures, cell-type counts correction, and spatial correlation analyses described above were applied identically to CIMS, with CIMS substituted as the exposure in place of COMET-predicted VAF.

## Supplementary Information

**Figure S1.**
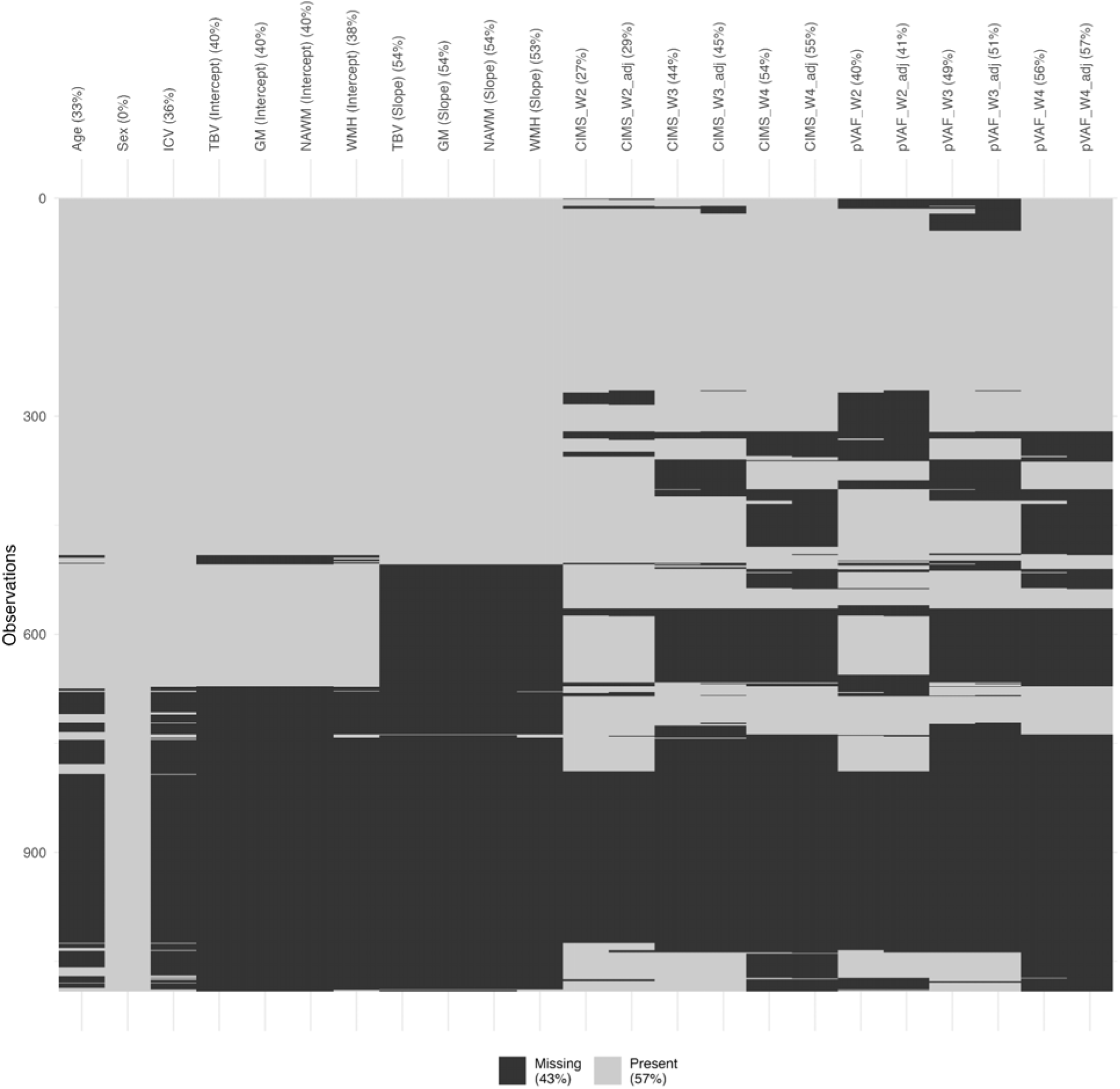
Missing data profile for the analytic dataset from the Lothian Birth Cohort 1936 (LBC1936). The plot visualises the proportion and clustering of missing values across all primary variables utilised in the latent growth curve models. Variables include demographic baseline covariates (age, sex, intracranial volume [ICV]), structural brain magnetic resonance imaging (MRI) metrics at baseline level (intercept) and their longitudinal rate of change (slope) for total brain volume (TBV), grey matter (GM), normal-appearing white matter (NAWM), and white matter hyperintensities (WMH). Longitudinal epigenetic predictors, COMET-predicted variant allele frequency (pVAF) and the EWAS-based CHIP-associated immune methylation score (CIMS), are shown across waves 2, 3, and 4, both before and after adjustment for leucocyte counts and technical batch. Hierarchical clustering (x-axis) groups variables with similar patterns of missingness.

**Figure S2.**
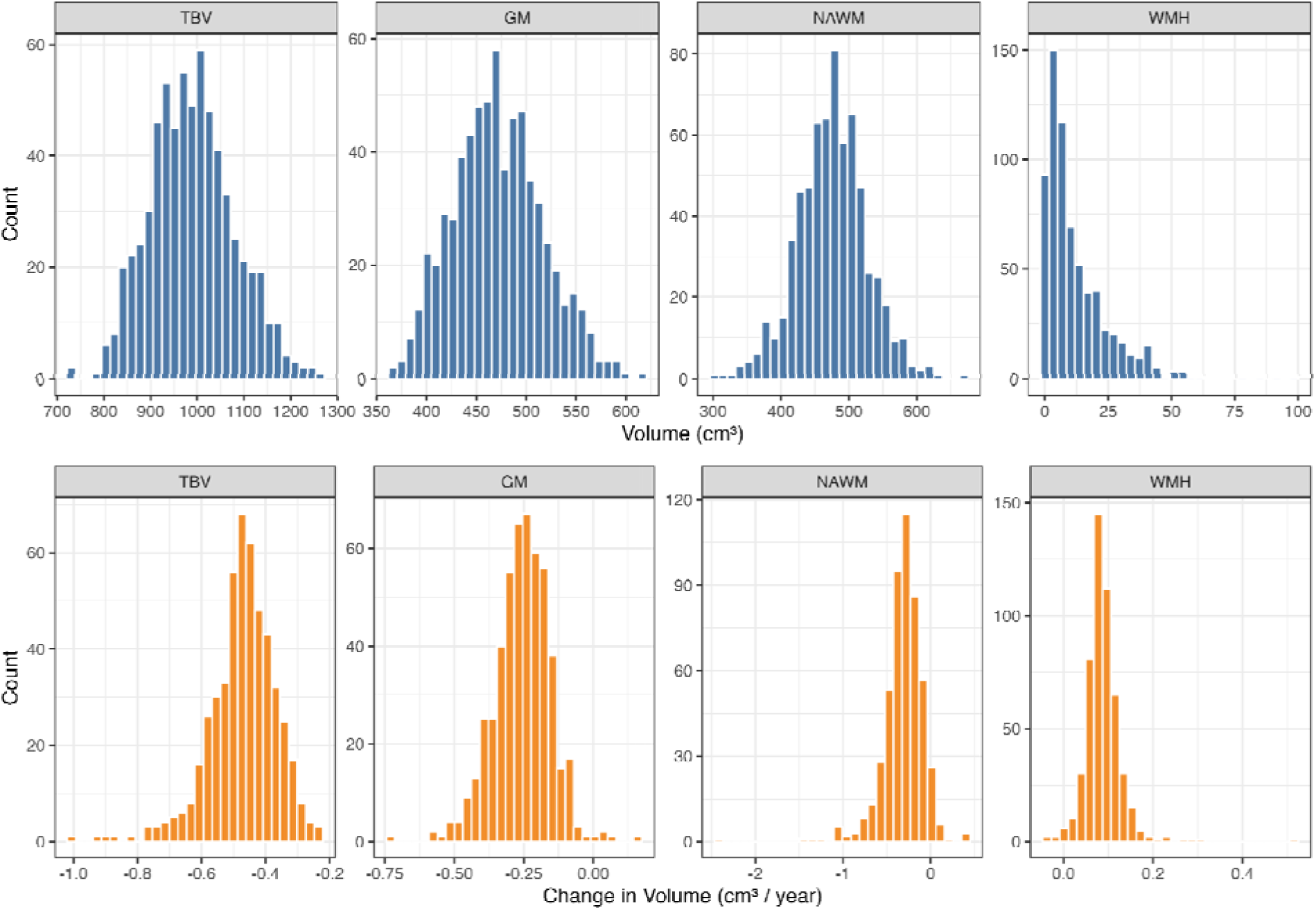
Distributions of baseline brain volumes and longitudinal rates of change. Histograms displaying the raw, unscaled structural magnetic resonance imaging (MRI) metrics for the Lothian Birth Cohort 1936 (LBC1936) sample. Top: Baseline volumes (model intercepts) assessed at wave 2, presented in cubic centimetres (cm³). Bottom: Longitudinal rate of volume change (model slopes) across the follow-up period, presented in cm³ per year (cm³/year). Measurements are stratified by tissue type: total brain volume (TBV), grey matter (GM), normal-appearing white matter (NAWM), and white matter hyperintensities (WMH). Negative slope values for TBV, GM, and NAWM indicate progressive tissue atrophy, whereas positive slope values for WMH denote progressive lesion expansion over time.

**Figure S3.**
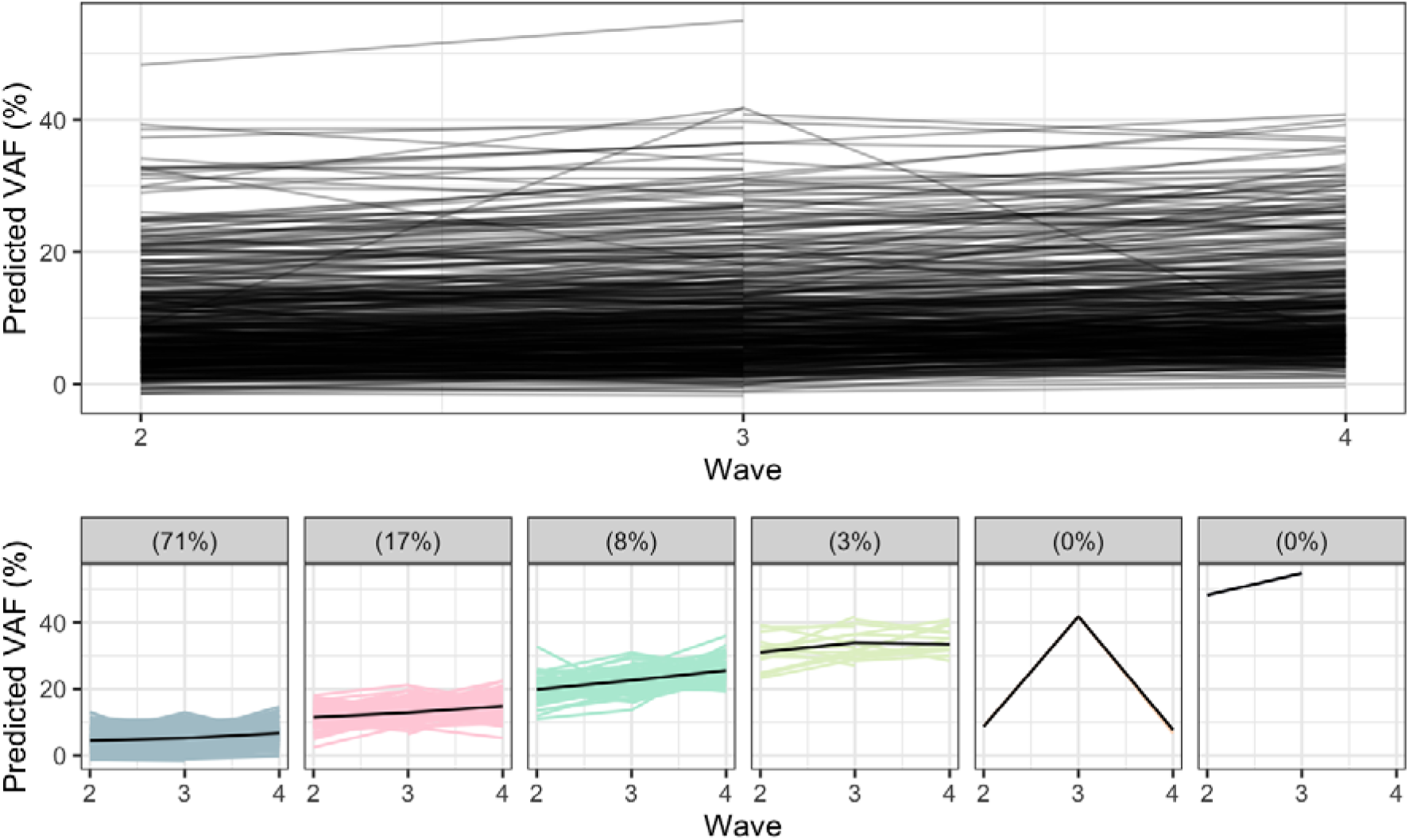
Longitudinal trajectories of COMET-predicted variant allele frequency (pVAF) across waves 2–4. Upper panel: Spaghetti plot demonstrating individual participant trajectories of pVAF over the follow-up period, illustrating cohort-wide trends in predicted clonal expansion. Lower panel: Unsupervised k-means for longitudinal data (KML) clustering reveals distinct sub-population trajectory profiles within the cohort. Each facet represents a unique cluster, highlighting the inter-individual heterogeneity in the baseline levels and progression rates of epigenetic CHIP signatures over time.

**Figure S4.**
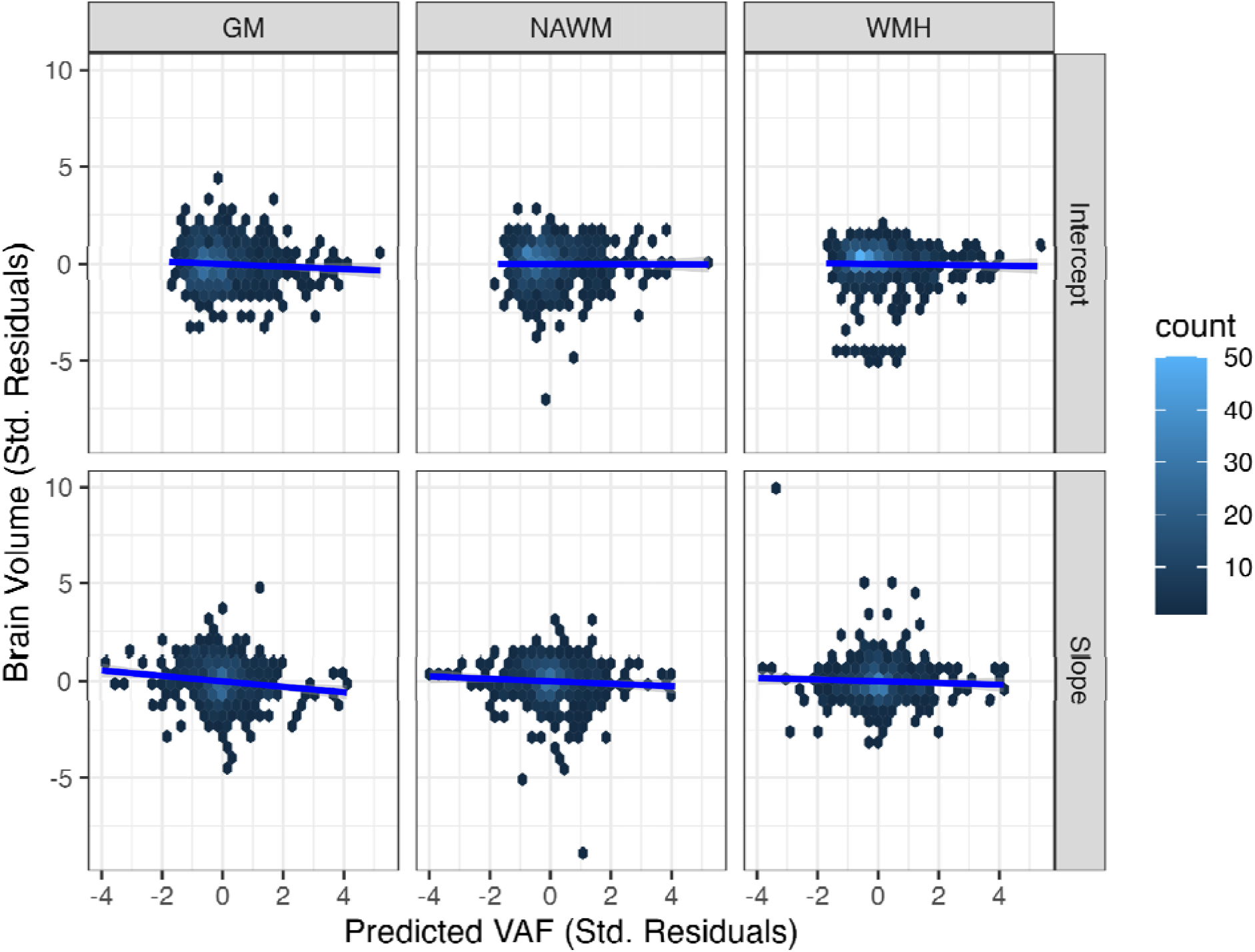
Partial correlations between COMET-predicted VAF and brain tissue-type measures at baseline level and rate of change. Scatter plots display the relationship between predicted variant allele frequency (pVAF) derived from COMET and brain tissue-type measures across three tissue types, grey matter (GM), normal-appearing white matter (NAWM), and white matter hyperintensities (WMH), stratified by the intercept (baseline level) and slope (rate of change) components of a latent growth curve model. Prior to plotting, VAF was regressed on age at MRI and sex, and each brain volume measure was regressed on age at MRI and sex; additionally for intercept [Top row], intracranial volume (ICV) was also included in the regression; standardised residuals from each model are displayed on the x- and y-axes, respectively, representing age- and sex-adjusted partial associations. Hex bins reflect local data density. Blue lines indicate ordinary least-squares fits with 95% confidence intervals. Both axes are expressed in standard deviation units to facilitate cross-panel comparison.

**Figure S5.**
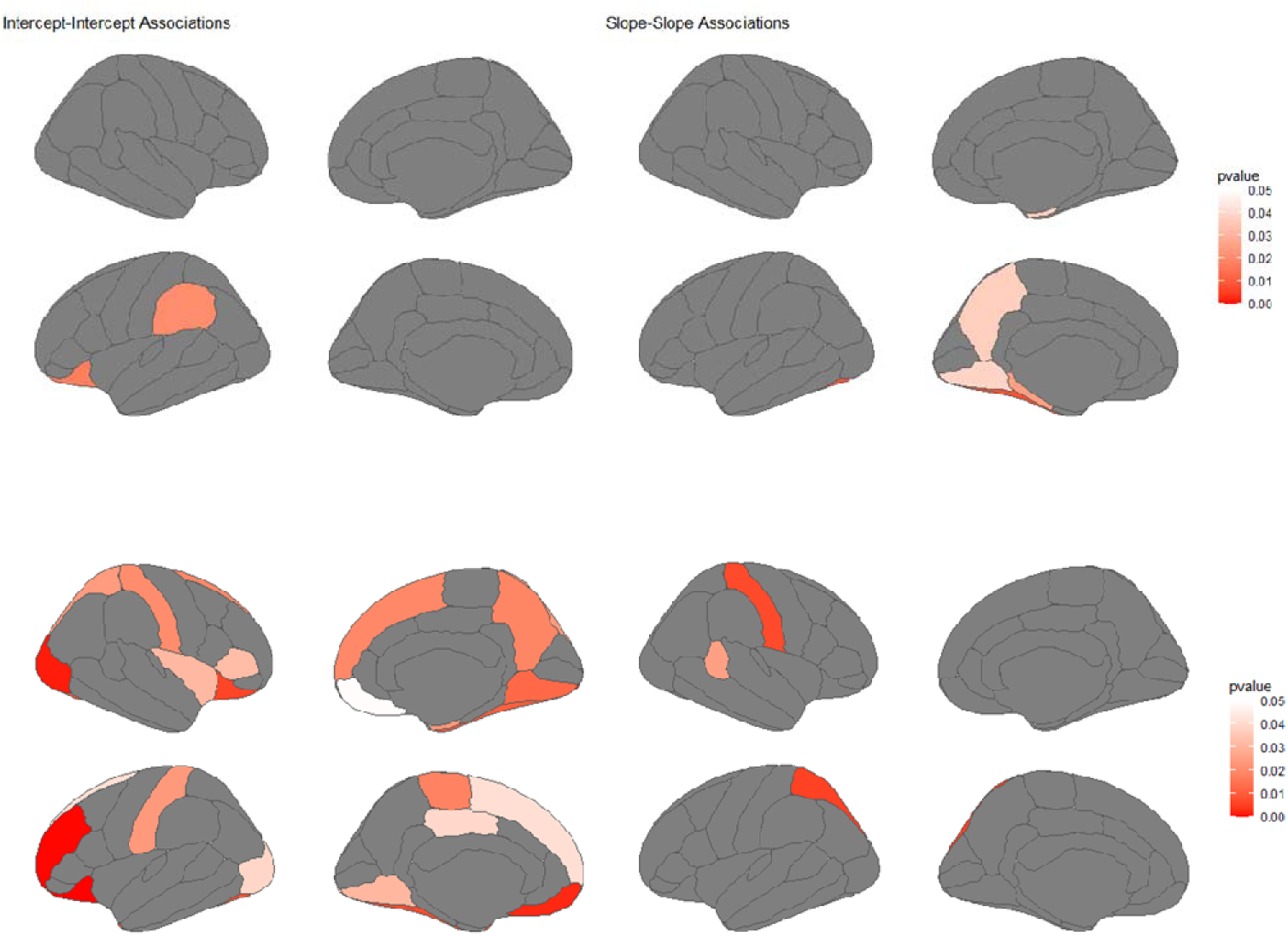
*p* values for associations between DNAm-based clonal haematopoiesis predictors and cortical volume at baseline (intercept–intercept) and rate of change (slope–slope). Latent growth curve modelling reveals region-specific associations between the two DNAm-based predictors and cortical volume across Desikan– Killiany (DK) parcellations, including cross-sectional associations (intercept–intercept) and longitudinal trajectory associations (slope–slope). Upper panel: results associated with COMET-derived pVAF. Lower panel: results associated with the EWAS-based CHIP-associated immune methylation score (CIMS). Darker red indicates lower (more significant) p-values. Grey regions did not pass the 0.05 significance threshold.

**Figure S6.**
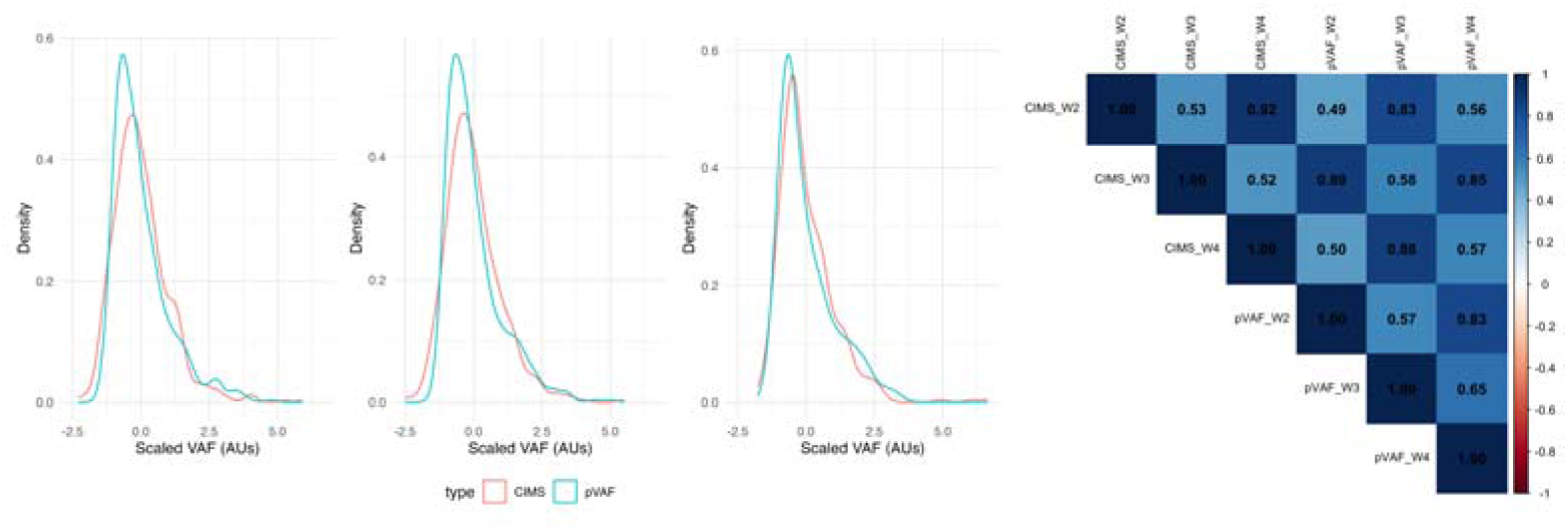
Characteristics of DNAm VAF predictors. **Left:** density plots of scaled VAF predictors (arbitrary units (AUs); M = 0, SD = 1) across measurement instances (which correspond to LBC1936 waves 2–4; ages 73, 76, 79). **Right:** heatmapped correlation matrix (Pearson *r*) across DNAm predictor types and measurement instances.

**Figure S7.**
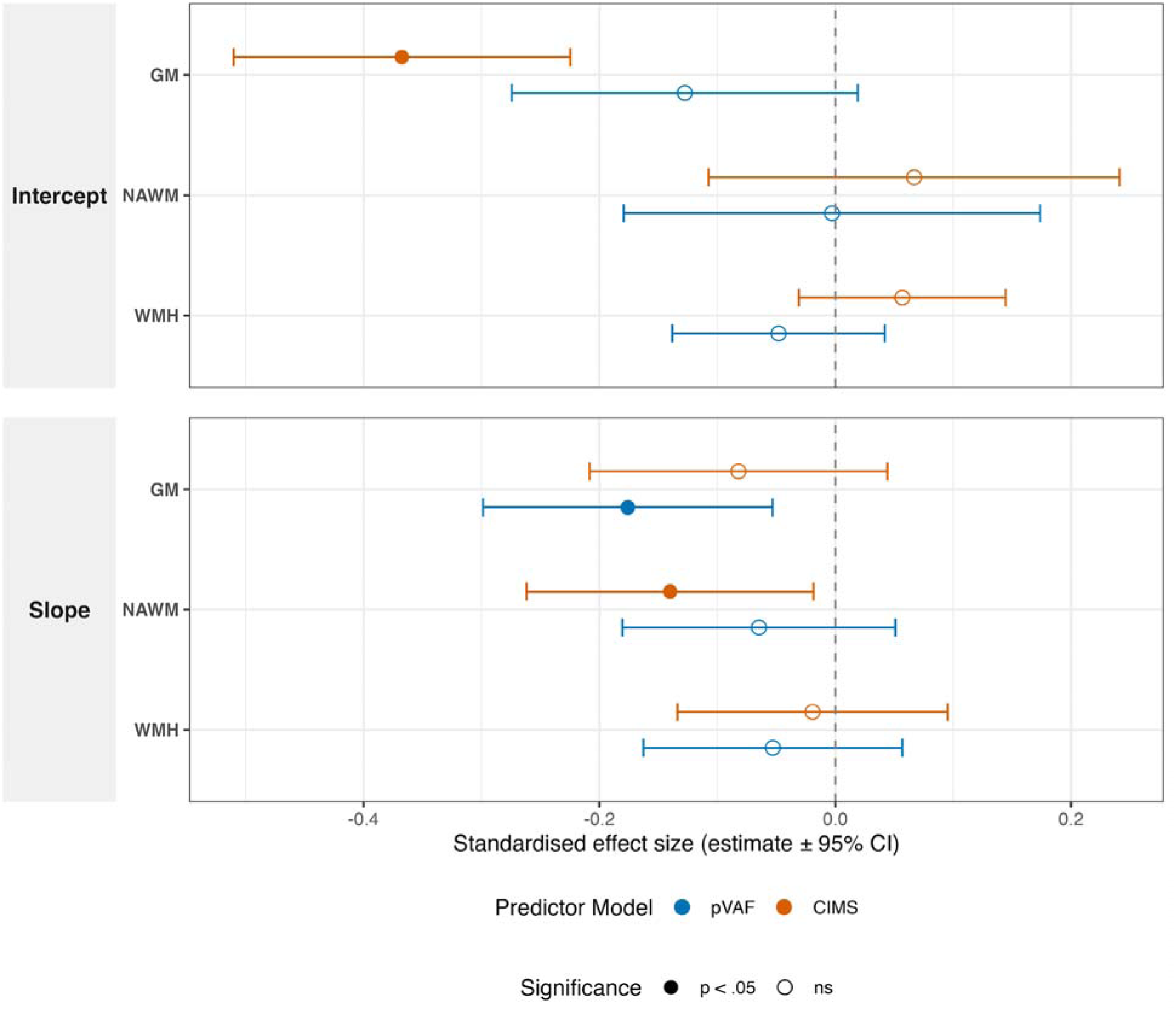
Standardised effect sizes for the association between DNA methylation-based CHIP predictors and longitudinal brain atrophy. Forest plot displaying the results of multivariable latent growth curve models adjusted for baseline age and sex. The plot compares the effect size of COMET-predicted VAF (pVAF; blue) and the EWAS based CHIP-associated immune methylation score (CIMS; orange). Standardised estimates are reported for both baseline brain volumes (Intercept, top panel) and longitudinal rates of volume change (Slope, bottom panel) across three tissue-type MRI measures: grey matter (GM), normal-appearing white matter (NAWM), and white matter hyperintensities (WMH). Error bars denote 95% confidence intervals. Solid points indicate statistically significant associations (*p* < 0.05), whereas hollow points denote non-significant findings.

**Figure S8.**
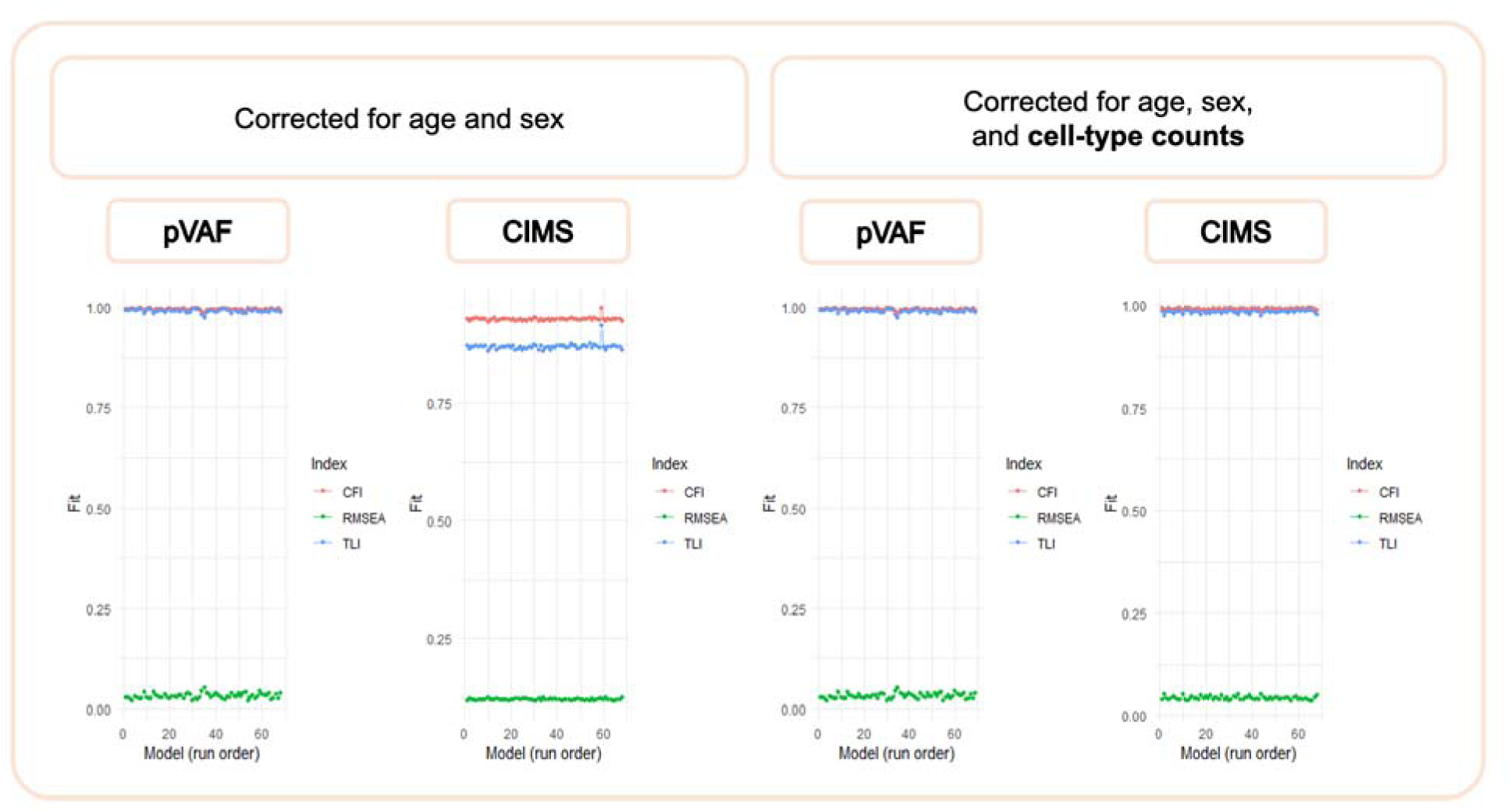
Model fit statistics across pVAF and CIMS models, before and after correction for cell-type counts. Fit indices (CFI, red; TLI, blue; RMSEA, green) are plotted for each model (x-axis, model run order) under two adjustment strategies: models corrected for age and sex only (left two panels), and models additionally corrected for cell-type counts (right two panels), shown separately for pVAF-based and CIMS-based models. CFI and TLI values close to 1.0 and RMSEA values close to 0 indicate good model fit. Across all conditions, models show consistently high CFI/TLI (∼0.9–1.0) and low RMSEA (∼0–0.1), indicating good fit.

## Contributors

Suwanlikit, Y. (Y.S.), Crofts, S.J.C. (S.J.C.C.), Cox, S.R. (S.R.C.), and Chandra, T. (T.C.) conceived and designed the study. S.J.C.C. performed the COMET-based clonality estimation. Bastin, M.E. (M.E.B.) and Wardlaw, J.M. (J.M.W.) secured funding for and oversaw acquisition of the original LBC structural MRI data. Moodie, J.E. (J.E.M.) and S.R.C. processed and quality-controlled the structural MRI data for this study. Marioni, R.E. (R.E.M.) developed the CIMS. Y.S., S.J.C.C., Robertson, N.A. (N.A.R.), and S.R.C. wrote the first draft of the manuscript. J.E.M., Tan, S.H. (S.H.T.), Kirschner, K. (K.K.), Vivithanaporn, P. (P.V.), M.E.B., and J.M.W. provided critical feedback on the data analysis and interpretation of results. S.R.C. and T.C. supervised the study and secured funding. All authors reviewed and edited the manuscript and approved the final version.

## Data Availability

Lothian Birth Cohort 1936 (LBC1936) data can be requested via a Data Request Form to the Lothian Birth Cohorts research team (https://lothian-birth-cohorts.ed.ac.uk/data-access-collaboration), subject to data access policies and governance procedures.

## Acknowledgements

We thank Dr. Baptiste Couvy-Duchesne for kindly providing the full set of regional dementia case-control effect-size estimates underlying the analyses reported in the meta-analytic dementia map study^44^, which were used in the spatial correlation analyses presented here. The Lothian Birth Cohorts were supported by joint funding from the BBSRC and ESRC (BB/W008793/1), Age UK (the Disconnected Mind project), the Medical Research Council (MRC; G0701120, G1001245, MR/M013111/1, and MR/R024065/1), the Milton Damerel Trust, and the University of Edinburgh. S.R.C. and J.E.M. were also supported by a Sir Henry Dale Fellowship, jointly funded by the Wellcome Trust and the Royal Society (221890/Z/20/Z). Y.S., S.J.C.C., N.A.R., S.H.T., K.K., and T.C. were funded by the Mayo Clinic Robert and Arlene Kogod Center on Aging. K.K. was funded by the Division of Hematology, Mayo Clinic, Rochester. J.M.W. was supported by the UK DRI funded by MRC, Alzheimer’s Society and ARUK. The funders had no role in study design, data collection, data analysis, data interpretation, or writing of the report.

## Ethics Approval

The LBC1936 study was given ethical approval by the Multi-Centre Research Ethics Committee for Scotland (MREC/01/0/56), the Lothian Research Ethics Committee (LREC/2003/2/29), and the Scotland A Research Ethics Committee (07/MRE00/58). All participants gave written informed consent.

## AI-Use Declaration

During preparation of this manuscript, the authors used AI tools, including but not limited to Claude and Gemini, solely to check grammar, spelling, and punctuation. All AI-assisted output was reviewed and edited by the authors, who take full responsibility for the accuracy and integrity of the manuscript.

## Research in context

### Evidence before this study

We searched PubMed for articles published from database inception to August 1, 2026, using the terms “clonal haematopoiesis”, “CHIP”, “clonal haematopoiesis of indeterminate potential”, “Alzheimer’s disease”, “dementia”, “cognitive impairment”, and “brain MRI”, without language restrictions. Clonal haematopoiesis of indeterminate potential (CHIP) is an established risk factor for cardiovascular disease, haematologic malignancy, and all-cause mortality, but its relationship with neurodegeneration is contradictory: some studies reported that CHIP was protective against cognitive impairment, while others identified it as a risk factor for stroke and Parkinson’s disease. Interestingly, the relationship with Alzheimer’s disease (AD) specifically is mixed, with reports of both protective and detrimental effects. Structural neuroimaging studies of CH have likewise been inconsistent: *DNMT3A*-mutant CHIP has been linked to lower white matter hyperintensity volume, suggesting a protective vascular effect, whereas another study found that CHIP overall, including *DNMT3A*-mutant CHIP, was linked to silent lesions, including microbleeds and white matter lesions. More importantly, this latter study was the only one to examine longitudinal imaging dynamics in relation to baseline CHIP status. More broadly, nearly all existing cohorts infer CH status from a single baseline blood draw, so no prior study has been able to track how changes in clonal burden relate to changes in brain structure over time. We identified no studies using repeated, methylation-based estimation of clonal burden alongside repeated structural MRI in the same individuals.

### Added value of this study

Using the Lothian Birth Cohort 1936, which uniquely combines blood DNA methylation and structural brain MRI collected at three timepoints over 6 years, we applied COMET, a mutation-agnostic, methylation-based predictor of clonal burden, together with latent growth curve modelling to separate baseline (cross-sectional) associations from longitudinal trajectories. Because COMET is mutation-agnostic, it captures the substantial proportion (∼78%) of clonal expansions that lack mutations in canonical driver genes and would be missed by targeted sequencing. To our knowledge, this is the first study to test whether longitudinal change in clonal haematopoiesis burden tracks longitudinal change in brain structure, rather than relying on a single static CH measurement. We found that clonality trajectories were associated with progressive, left-lateralised cortical atrophy in medial temporal and parietal regions that statistically significantly overlapped with an established AD atrophy signature, while showing nominal relative preservation of the caudate and no significant hippocampal association, a pattern that only partially recapitulates canonical AD topography. We further developed CIMS, a complementary methylation score derived from CHIP epigenome-wide association study summary statistics, and found that it converged with COMET on global brain atrophy but diverged regionally, indicating that clone-size-based and broader epigenomic-remodelling-based measures capture distinct, non-redundant aspects of CH biology.

### Implications of all the available evidence

These findings suggest that blood-based clonal haematopoiesis burden, assessed longitudinally, is associated with progressive AD-like patterns of cortical atrophy in community-dwelling older adults, adding to the body of evidence supporting CH burden as a candidate blood-based marker of age-related neurodegeneration. The divergence from canonical AD subcortical involvement, together with the two only partly overlapping methylation-based signals (COMET and CIMS), suggests CH-related brain change may reflect a partially distinct biological route rather than a direct recapitulation of AD pathology. Replication in independent, ideally more diverse cohorts, integration with mutation-level sequencing, and pairing with amyloid/tau biomarkers are needed to determine whether clonal burden marks AD pathology directly, a parallel pathway of cortical vulnerability, or accelerated normative ageing.

