## Supplementary material for "Clonal Haematopoiesis Tracks Alzheimer’s-like Cortical Atrophy in Community-Dwelling Older Adults": Figure S

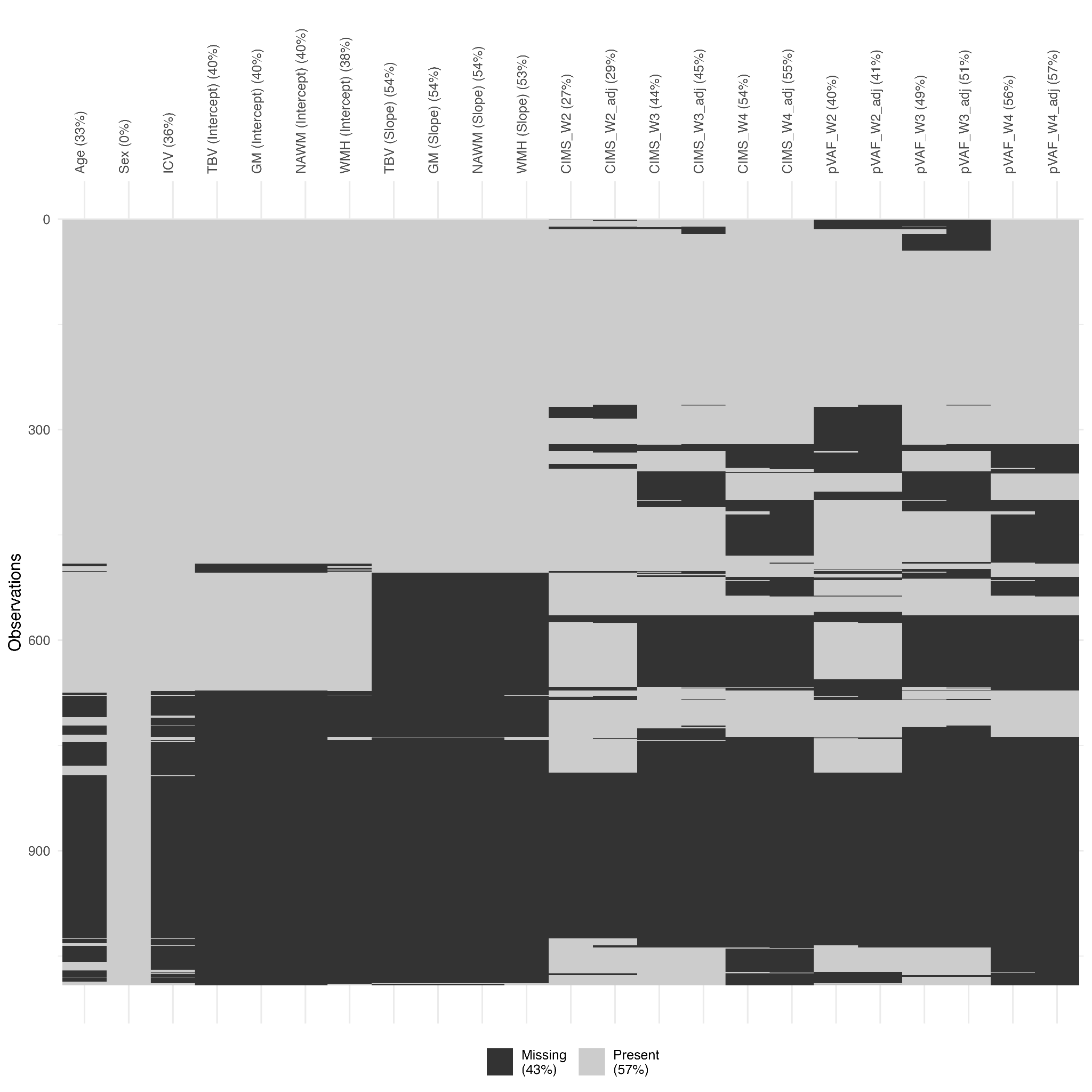

**Figure S1. Missing data profile for the analytic dataset from the Lothian Birth Cohort 1936 (LBC1936).** The plot visualises the proportion and clustering of missing values across all primary variables utilised in the latent growth curve models. Variables include demographic baseline covariates (age, sex, intracranial volume [ICV]), structural brain magnetic resonance imaging (MRI) metrics at baseline level (intercept) and their longitudinal rate of change (slope) for total brain volume (TBV), grey matter (GM), normal-appearing white matter (NAWM), and white matter hyperintensities (WMH). Longitudinal epigenetic predictors, COMET-predicted variant allele frequency (pVAF) and the EWAS-based CHIP-associated immune methylation score (CIMS), are shown across waves 2, 3, and 4, both before and after adjustment for leucocyte counts and technical batch. Hierarchical clustering (x-axis) groups variables with similar patterns of missingness.

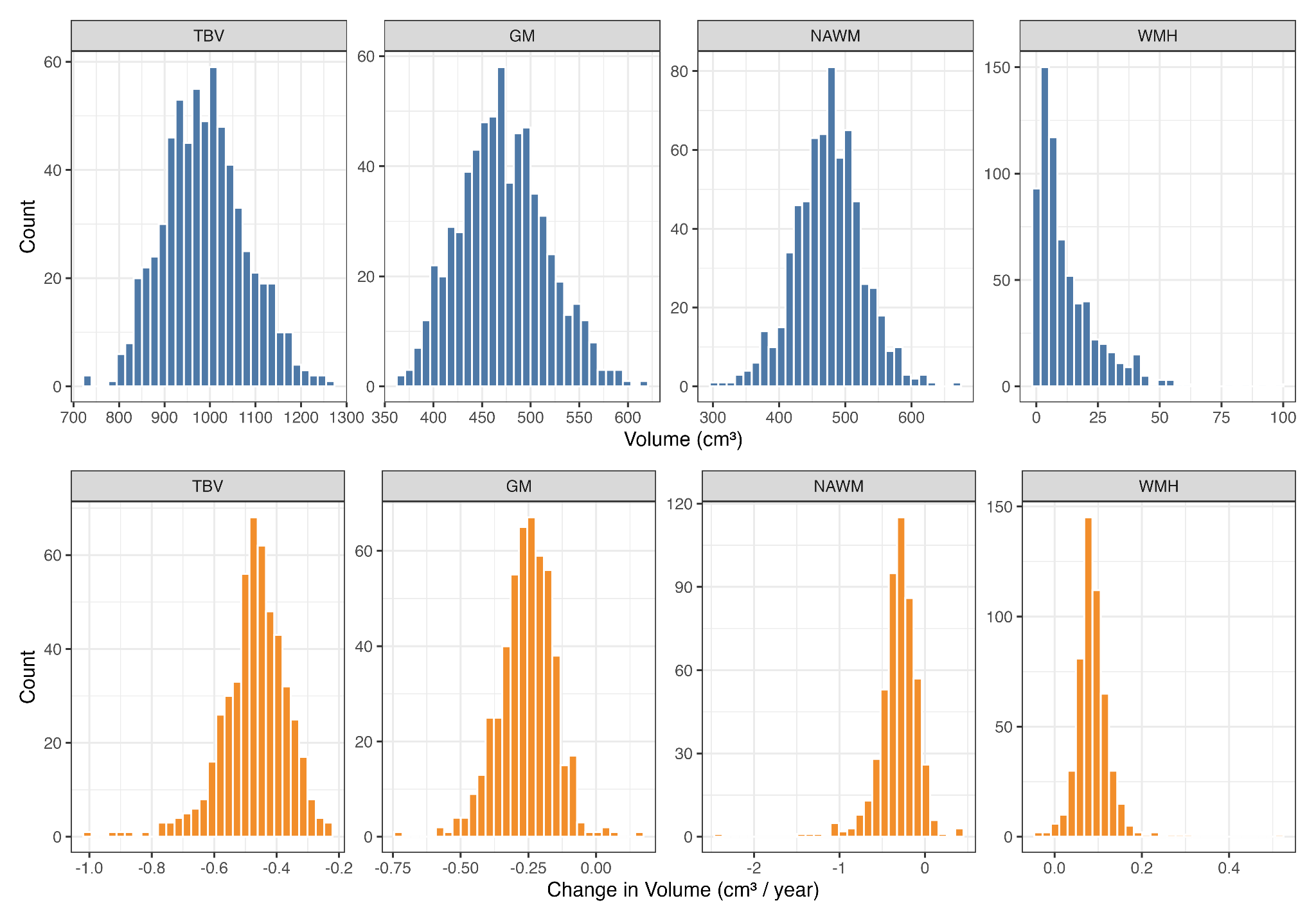

**Figure S2. Distributions of baseline brain volumes and longitudinal rates of change.** Histograms displaying the raw, unscaled structural magnetic resonance imaging (MRI) metrics for the Lothian Birth Cohort 1936 (LBC1936) sample. Top: Baseline volumes (model intercepts) assessed at wave 2, presented in cubic centimetres (cm³). Bottom: Longitudinal rate of volume change (model slopes) across the follow-up period, presented in cm³ per year (cm³/year). Measurements are stratified by tissue type: total brain volume (TBV), grey matter (GM), normal-appearing white matter (NAWM), and white matter hyperintensities (WMH). Negative slope values for TBV, GM, and NAWM indicate progressive tissue atrophy, whereas positive slope values for WMH denote progressive lesion expansion over time.

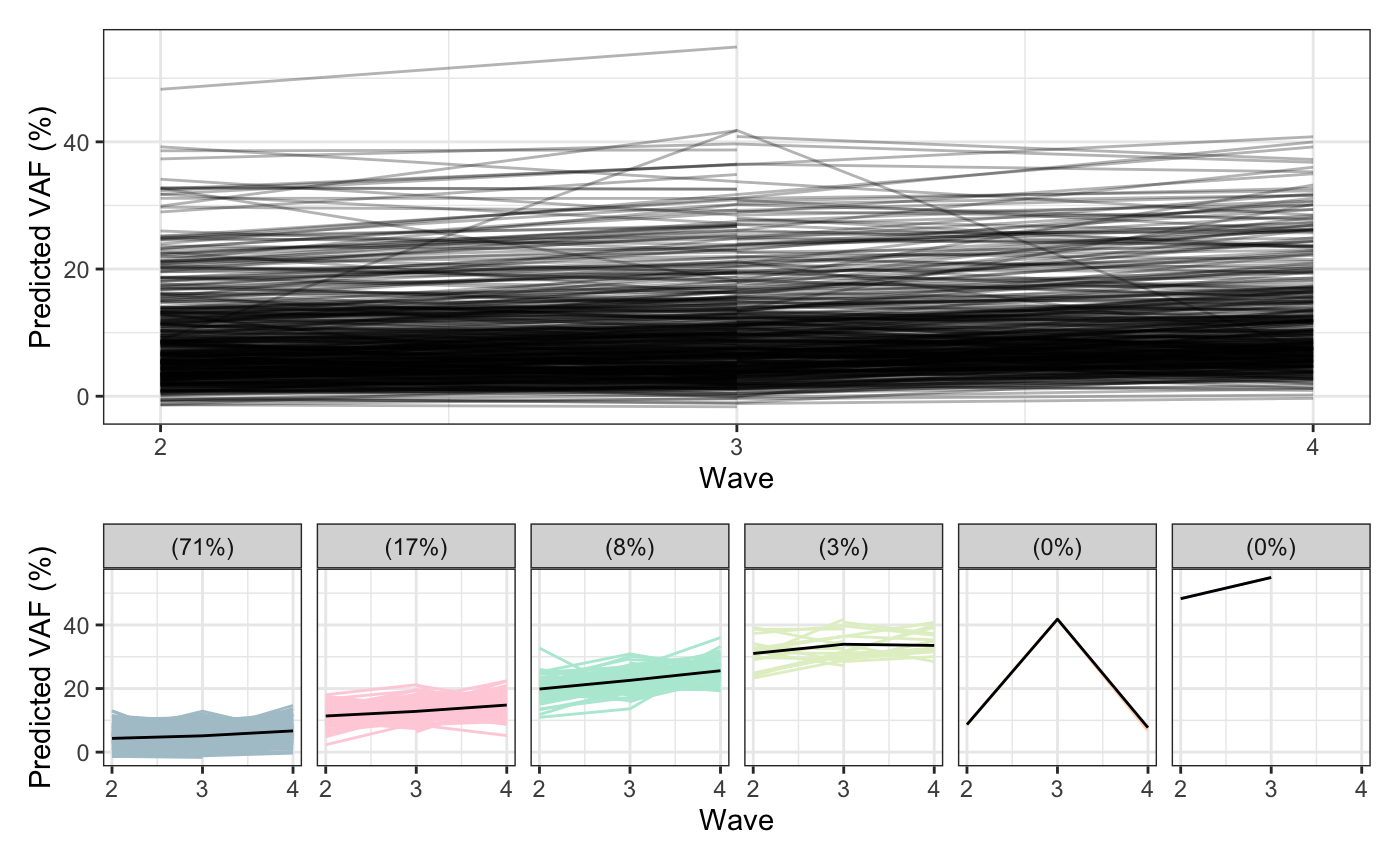

**Figure S3. Longitudinal trajectories of COMET-predicted variant allele frequency (pVAF) across waves 2–4**. Upper panel: Spaghetti plot demonstrating individual participant trajectories of pVAF over the follow-up period, illustrating cohort-wide trends in predicted clonal expansion. Lower panel: Unsupervised k-means for longitudinal data (KML) clustering reveals distinct sub-population trajectory profiles within the cohort. Each facet represents a unique cluster, highlighting the inter-individual heterogeneity in the baseline levels and progression rates of epigenetic CHIP signatures over time.

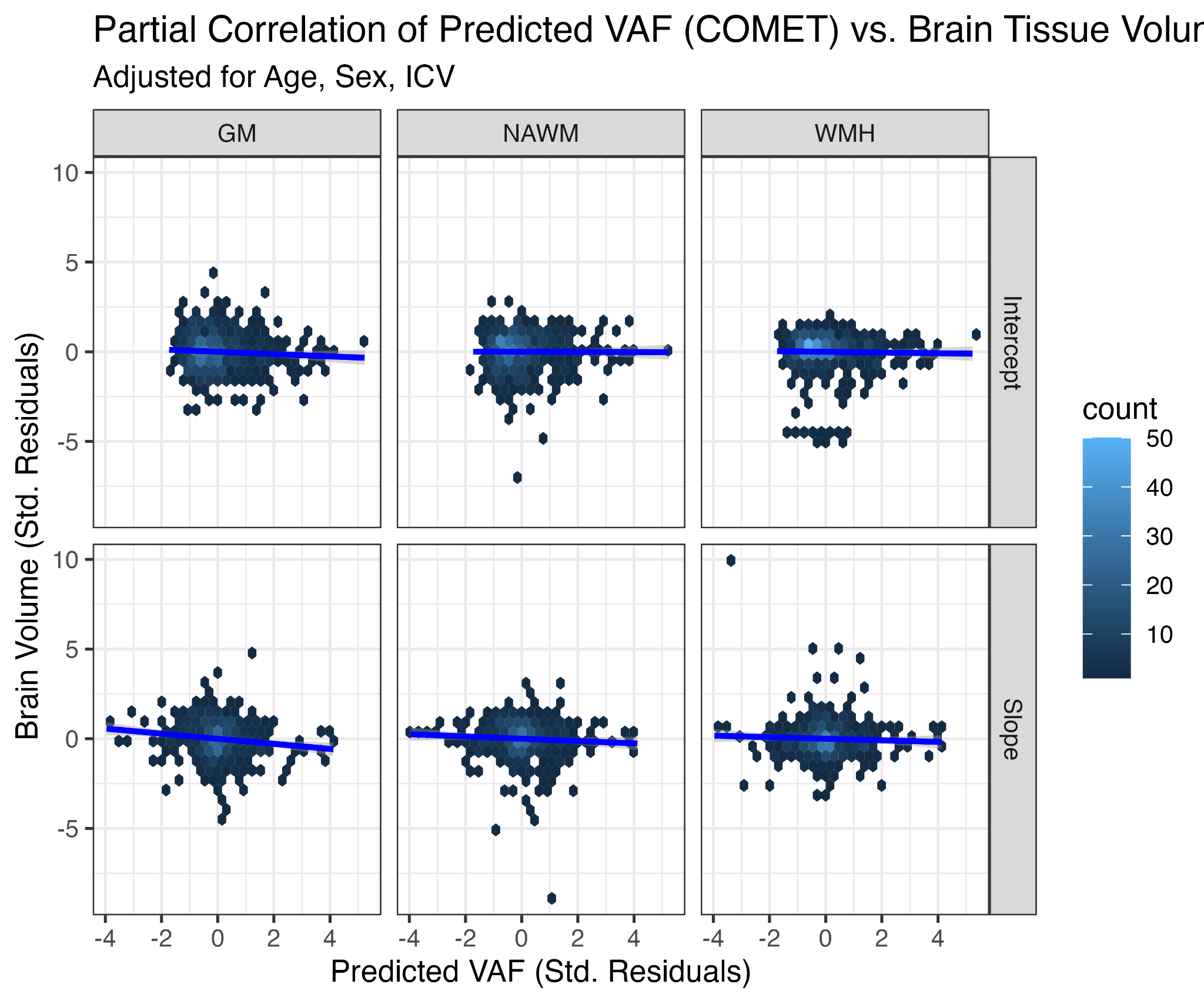

**Figure S4. Partial correlations between COMET-predicted VAF and brain tissue-type measures at baseline level and rate of change.** Scatter plots display the relationship between predicted variant allele frequency (pVAF) derived from COMET and brain tissue-type measures across three tissue types, grey matter (GM), normal-appearing white matter (NAWM), and white matter hyperintensities (WMH), stratified by the intercept (baseline level) and slope (rate of change) components of a latent growth curve model. Prior to plotting, VAF was regressed on age at MRI and sex, and each brain volume measure was regressed on age at MRI, sex; additionally for intercept [Top row], intracranial volume (ICV) was also included in the regression; standardised residuals from each model are displayed on the x- and y-axes, respectively, representing age- and sex-adjusted partial associations. Hex bins reflect local data density. Blue lines indicate ordinary least-squares fits with 95% confidence intervals. Both axes are expressed in standard deviation units to facilitate cross-panel comparison.

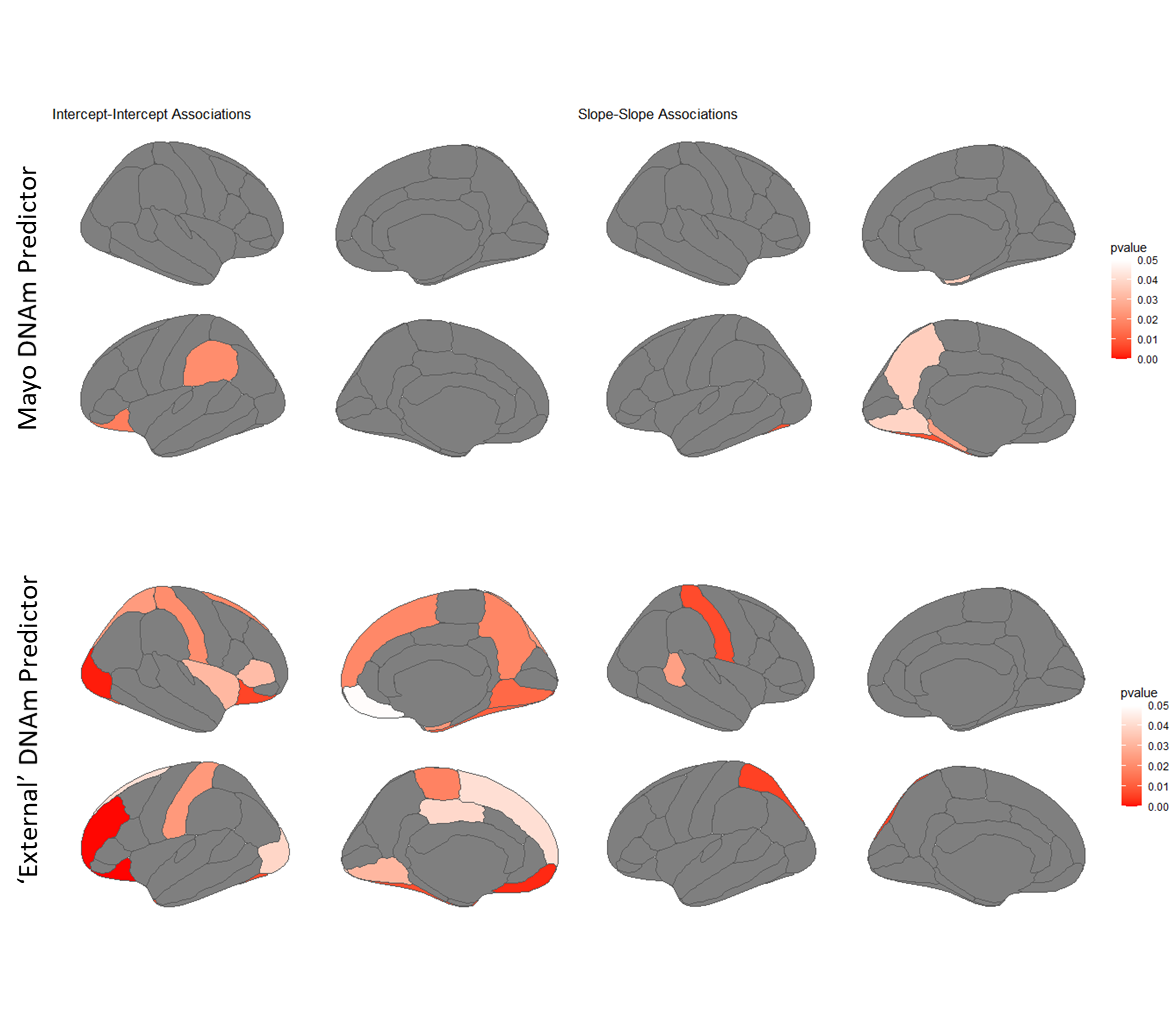

**Figure S5. *p* values for associations between DNAm-based clonal haematopoiesis predictors and cortical volume at baseline (intercept–intercept) and rate of change (slope–slope).** Latent growth curve modelling reveals region-specific associations between the two DNAm-based predictors and cortical volume across Desikan–Killiany (DK) parcellations, including cross-sectional associations (intercept–intercept) and longitudinal trajectory associations (slope–slope). Upper panel: results associated with COMET-derived pVAF. Lower panel: results associated with the EWAS-based CHIP-associated immune methylation score (CIMS). Darker red indicates lower (more significant) p-values. Grey regions did not pass the 0.05 significance threshold.

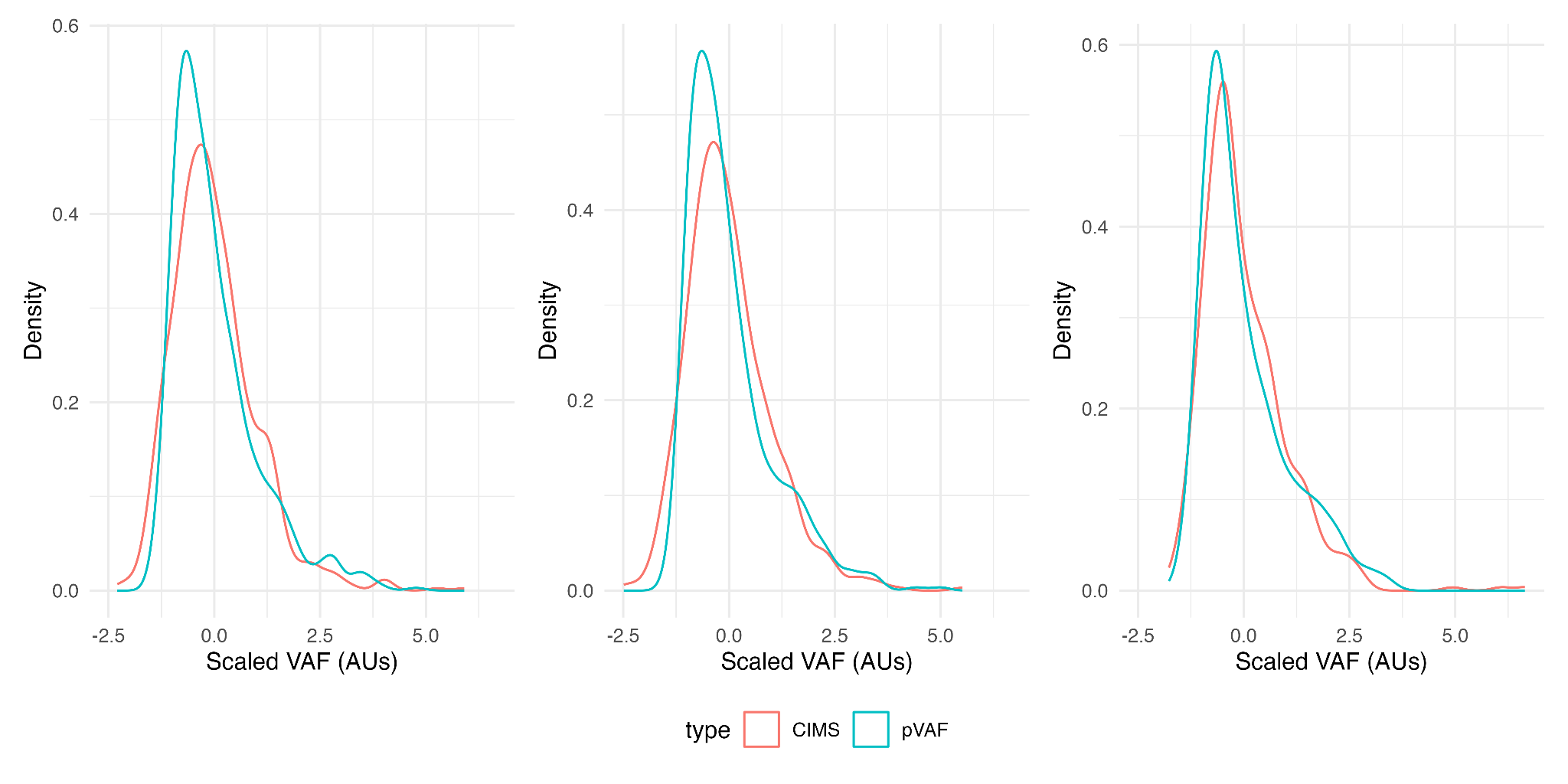

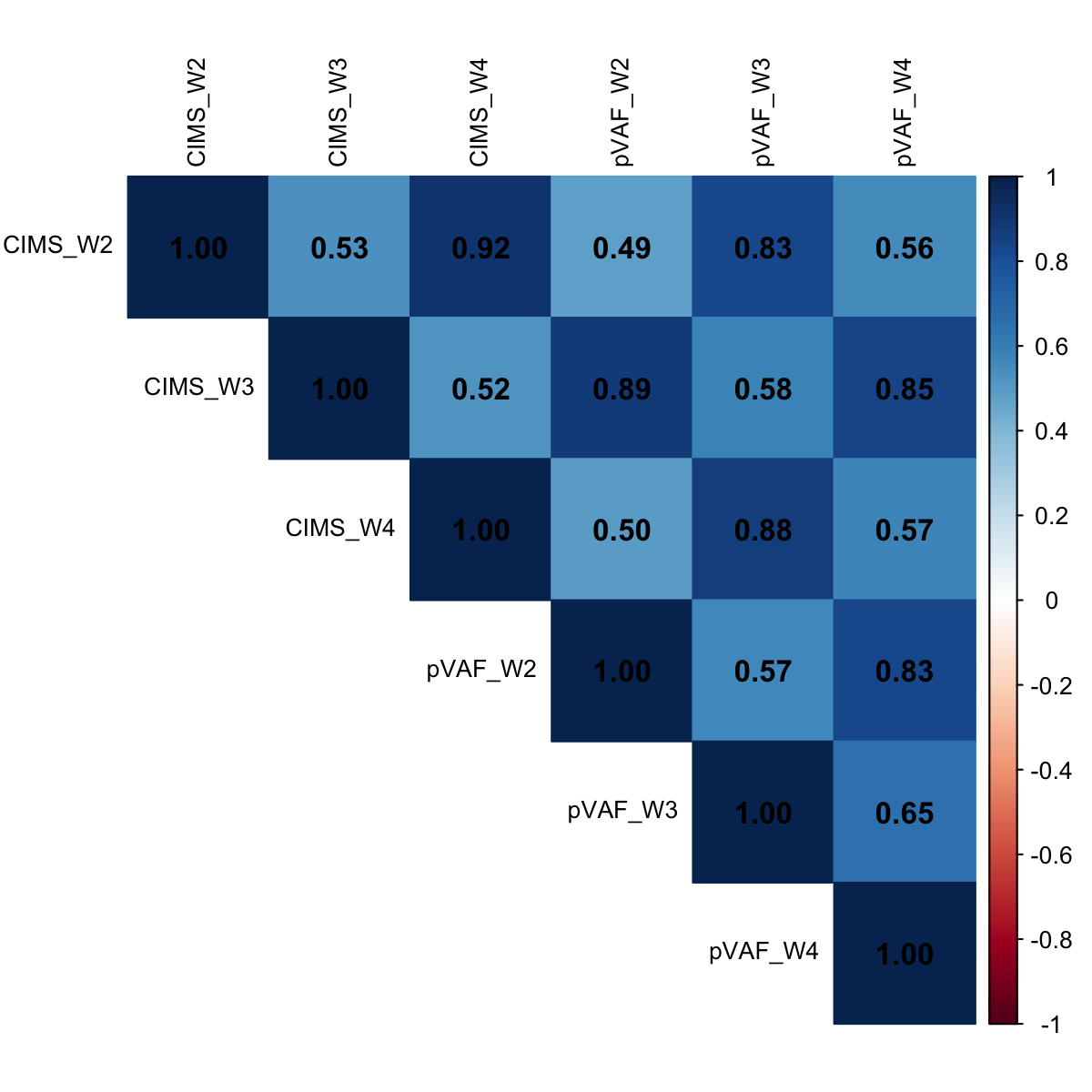

**Figure S6. Characteristics of DNAm VAF predictors**. **Left:** density plots of scaled VAF predictors (arbitrary units (AUs); M = 0, SD = 1) across measurement instances (which correspond to LBC1936 waves 2–4; ages 73, 76, 79). **Right:** heatmapped correlation matrix (Pearson *r*) across DNAm predictor types and measurement instances.

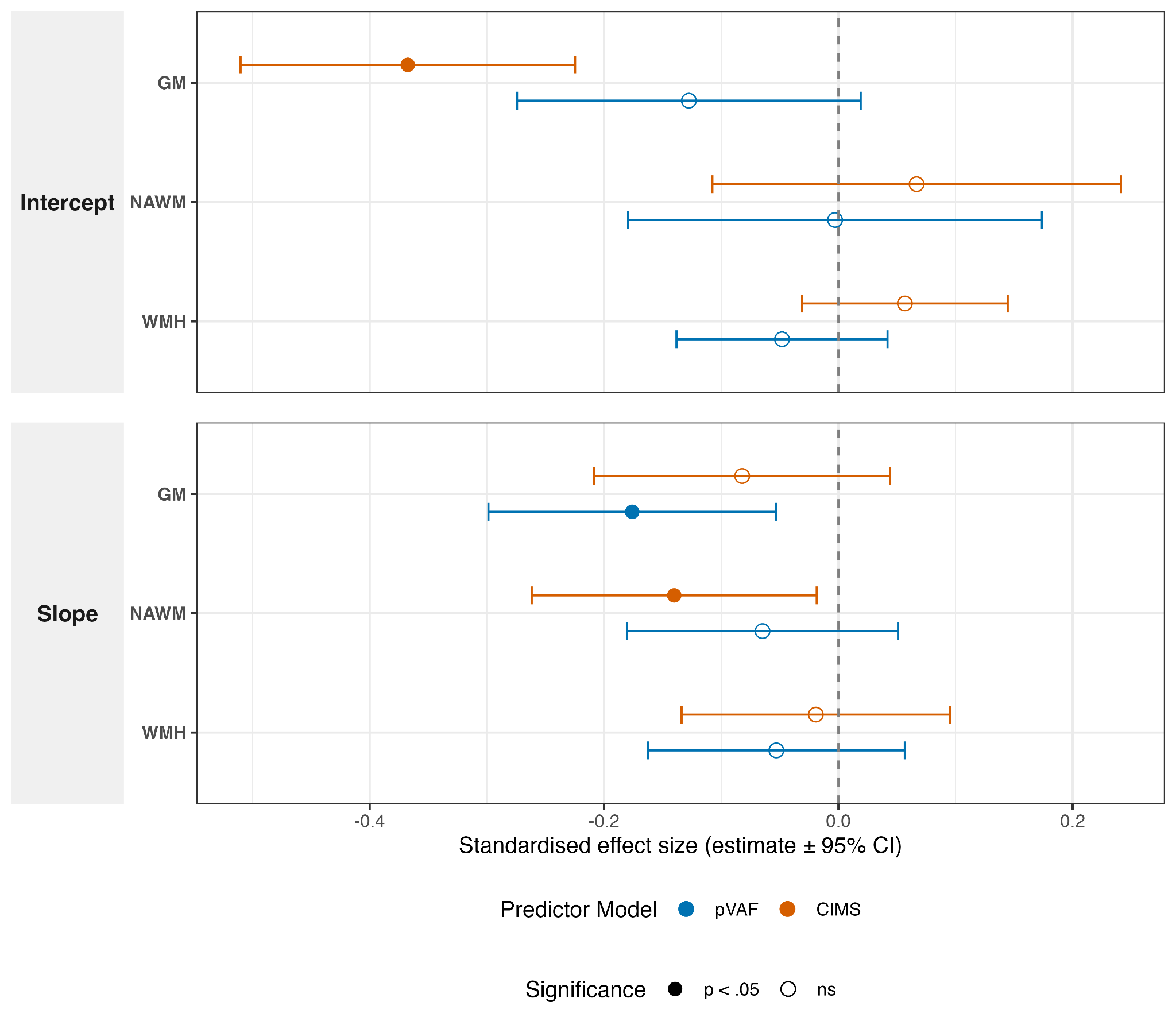

**Figure S7. Standardised effect sizes for the association between DNA methylation-based CHIP predictors and longitudinal brain atrophy.** Forest plot displaying the results of multivariable latent growth curve models adjusted for baseline age, sex. The plot compares the effect size of COMET-predicted VAF (pVAF; blue) and the EWAS-based CHIP-associated immune methylation score (CIMS; orange). Standardised estimates are reported for both baseline brain volumes (Intercept, top panel) and longitudinal rates of volume change (Slope, bottom panel) across three tissue-type MRI measures: grey matter (GM), normal-appearing white matter (NAWM), and white matter hyperintensities (WMH). Error bars denote 95% confidence intervals. Solid points indicate statistically significant associations (*p* < 0.05), whereas hollow points denote non-significant findings.

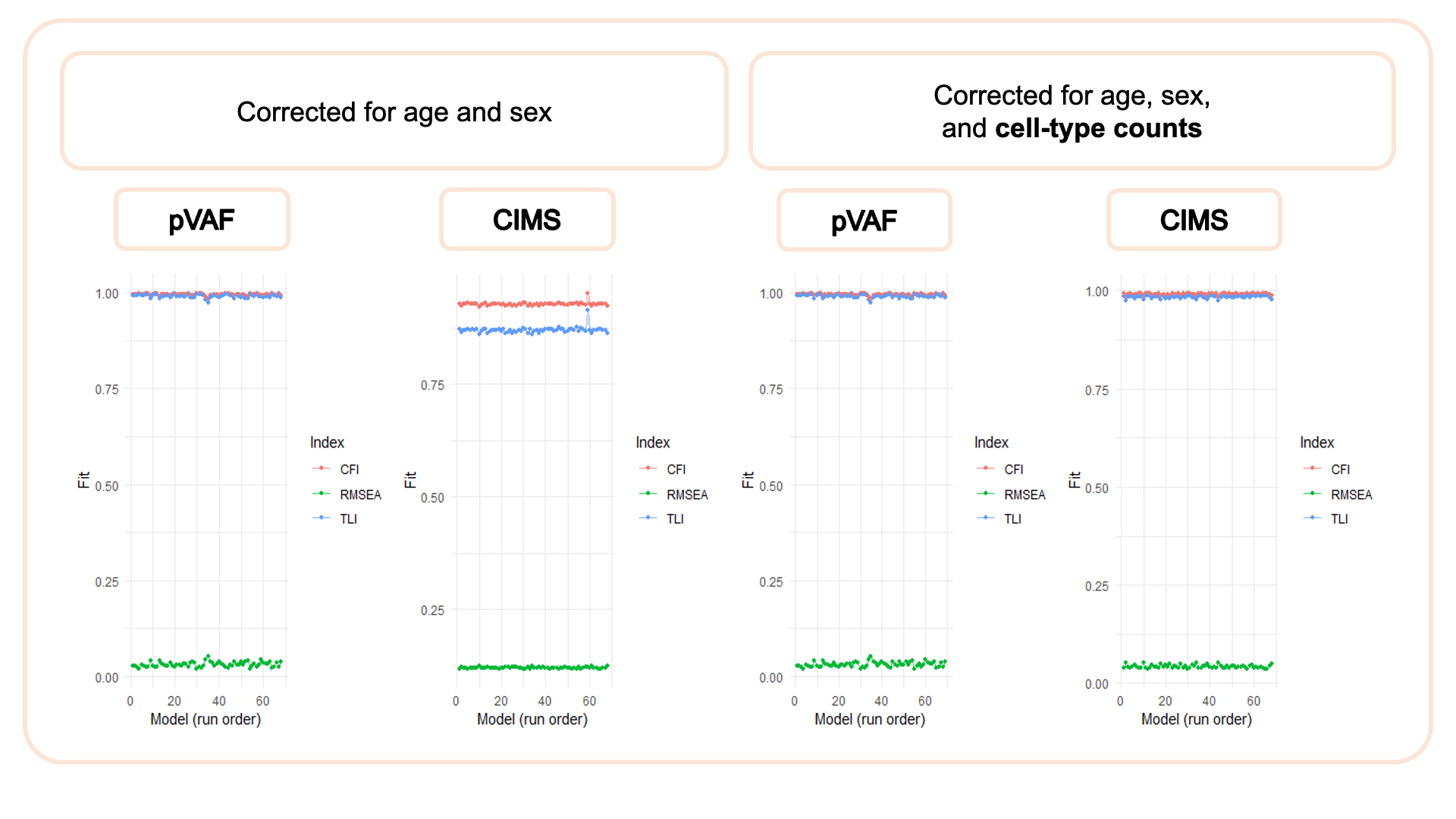

**Figure S8. Model fit statistics across pVAF and CIMS models, before and after correction for cell-type counts.** Fit indices (CFI, red; TLI, blue; RMSEA, green) are plotted for each model (x-axis, model run order) under two adjustment strategies: models corrected for age and sex only (left two panels), and models additionally corrected for cell-type counts (right two panels), shown separately for pVAF-based and CIMS-based models. CFI and TLI values close to 1.0 and RMSEA values close to 0 indicate good model fit. Across all conditions, models show consistently high CFI/TLI (~0.9–1.0) and low RMSEA (~0–0.1), indicating good fit.
